# Co-designing and optimising a primary care implementation model for childhood pneumonia management in a low-resource setting using principles of implementation research

**DOI:** 10.64898/2026.07.16.26358238

**Authors:** Barsha Gadapani Pathak, Sarmila Mazumder, Aditya Bhatt, Moammar Hashmi, Mandeep Singh, Tarun Madhur, Virender Yadav, Suresh Bhonsle, Rahul Sharma, Yasir Bin Nisar, Ingvild Fossgard Sandøy

**Affiliations:** Society for Applied Studies, New Delhi, India; Centre for Intervention Science in Maternal and Child Health, Centre for International Health, Department of Global Public Health and Primary Care, University of Bergen, Bergen, Norway; Maternal and Child Health, National Health Mission, Chandigarh, Haryana, India; Palwal District Health Authority, Palwal, Haryana, India; Department of Maternal, Newborn, Child and Adolescent Health and Ageing (MCA), World Health Organization (WHO), Geneva, Switzerland

## Abstract

**Background:** Pneumonia remains the leading infectious cause of under-five mortality, particularly in low- and middle-income countries (LMICs) where translation of evidence-based guidelines into practice faces systemic barriers. Although India launched the Childhood Pneumonia Management Guidelines (CPMG), effective implementation remains suboptimal. This study demonstrates how implementation research can support the systematic development and optimisation of a primary care--focused implementation model for childhood pneumonia management in a low-resource setting.

**Methods:** This manuscript reports findings from Phases I and II of a 26-month, pre-post quasi-experimental implementation research project conducted in Palwal district, Haryana, India. The Phase I is formative research including a baseline survey (June-August 2023); Phase II: development and iterative optimisation of the implementation model through co-design within a learning cluster (September 2023-May 2024); and Phase III: implementation and evaluation of the optimised model followed by district-wide scale-up (May 2024-March 2025). Phase I (June-August 2023) employed the Consolidated Framework for Implementation Research (CFIR) to systematically identify multilevel determinants affecting CPMG implementation across a study area of approximately 108,000 inhabitants. Phase II (September 2023-May 2024) focused on co-designing context-specific implementation strategies using the CFIR-ERIC (Consolidated Framework for Implementation Research-Expert Recommendations for Implementing Change) Barrier Busting Tool and iteratively refining an implementation model through participatory co-design workshops with government and community stakeholders within a learning block of approximately 50,000 inhabitants. An optimised Implementation Research Logic Model (IRLM) was developed to align determinants, strategies, mechanisms of change, and implementation outcomes.

**Results:** Thirty-three tailored implementation strategies were identified, addressing barriers across inner (e.g. implementation climate), outer (e.g., socio-cultural norms), individual (e.g. skills), process (e.g., planning), and innovation (e.g., complexity) CFIR domains. Iterative refinement through three successive models (Model 0+, Model 1, and Model 2) within the learning block was associated with improvements in key outcomes: care-seeking from appropriate primary care facilities increased from 0.8% at baseline to over 76% [193/254 cases], appropriate diagnosis improved from 0% to 92.7% [179/193 cases], and fidelity to guideline-based management reached 86% [154/179 cases] by the end of Phase II. These improvements informed the finalised IRLM which was subsequently implemented and evaluated in Phase-III.

**Conclusions:** This study demonstrates a systematic, transparent, and participatory approach to developing and optimising an implementation model for primary care-based childhood pneumonia management in a resource-constrained setting. The co-designed IRLM, grounded in established implementation research frameworks, offers a replicable process for translating evidence-based guidelines into primary care practice in similar LMIC settings. Phase III along with the district-wide implementation and evaluation findings will be reported in a forthcoming paper.

**Trial Registration:** Clinical trial registry CTRI/2021/03/031622 [date: 01/03/2021].

## INTRODUCTION

Despite advancements in healthcare and a reduction in global under-five mortality, pneumonia remains the leading infectious cause of death among children in this age group.[1] Globally, lower respiratory tract infections, including pneumonia, resulted in an estimated 2.18 million deaths in 2021, primarily among children under five years of age and adults over 70 years. [2] Pneumonia accounted for over 800,000 deaths worldwide and the highest burden in low- and middle-income countries (LMICs), where access to quality healthcare remains inadequate.[3] In India, approximately 30 million cases of pneumonia are reported annually, and the disease accounts for 15% of all under-five deaths against an under-five mortality rate of 41.9 per 1,000 live births.[4-6]

Recognizing the importance of early identification and prompt treatment at the closest level of care, the Indian government introduced the Childhood Pneumonia Management Guidelines (CPMG) alongside the Social Awareness and Action to Neutralize Pneumonia (SAANS) campaign in 2019.[7] Under the CPMG and SAANS initiative, Health and Wellness Centres (HWCs) were explicitly envisaged as the primary hub for pneumonia prevention, detection, and outpatient management, representing the most peripheral and accessible units of India’s primary health-care system.[8] HWCs, established under the Ayushman Bharat programme, are designed to deliver comprehensive primary care services, including child health, at the community level, thereby serving as the first point of contact for caregivers and under-five children. [9, 10] The CPMG emphasizes OPD-based management of non-severe pneumonia at primary care facilities, including HWCs and Primary Health Centres (PHCs), with timely referral of severe cases to higher-level facilities.[8] This primary care-based approach aims to strengthen pneumonia prevention, enable timely case detection and appropriate referral, improve adherence to management protocols, and support capacity-building of healthcare workers through training.[8] Collectively, these measures are intended to improve caregiver accessibility and treatment compliance while reducing avoidable burden on secondary and tertiary health facilities.[7]

In November 2022, Haryana, a North Indian state, launched its first SAANS campaign, initiating the roll-out of the CPMG across public-sector facilities. [11] The implementation was delayed by disruptions related to the COVID-19 pandemic.[11] As seen in many LMIC settings, translating evidence-based clinical guidelines into routine primary care practice remains challenging due to systemic constraints, including limited infrastructure, workforce shortages, inconsistent medicine supply, weak information systems, and community-level barriers to care-seeking.[9, 10, 12] These challenges are often most pronounced at HWCs, where frontline providers are expected to deliver guideline-based outpatient care while concurrently implementing multiple national health programmes.[9, 10] Implementation research offers a structured approach to understanding and addressing such challenges by examining how evidence-based interventions are adopted, implemented, and sustained in real-world primary care settings.[13]

To address these implementation challenges, a three-phase implementation research project was under-taken in Palwal district, Haryana. The following summarises previously published findings from Phase I of this project, using implementation research principles, which are included here as contextual background and are not reproduced as new results in this paper. [11, 14,15] The formative phase I of the study (conducted in June-August 2023) systematically identified multi-level implementation determinants using CFIR framework.[14] Supply-side barriers including infrastructure gaps, privacy concerns in health and wellness centres (HWCs), medicine supply disruptions, workforce training deficit, shortages of functional equipment and weak health information systems. [11] Demand-side barriers including caregivers y reluctance to seek care from government facilities, preference for informal healthcare providers, and low awareness of pneumonia dangers signs- are reported in full in two companion publications.[11, 15] In summary, Phase-I revealed that the CPMG had not been implemented in Palwal prior to study initiation, establishing a true baseline context for model development.[11, 15] Key facilitators identified included problem-solving capacity among some frontline providers, availability of untied funds at HWCs and PHCs, and potential for community engagement.[11] These findings directly informed Phase II, which is the primary focus of this paper. The Phase II of the implementation research focused on designing contextually relevant implementation strategies and developing an optimized model to strengthen pneumonia management at primary level of health care. Although primary level of healthcare facilities, namely HWCs and PHCs, were the central focus of implementation, Community Health Centres (CHCs) and the District Hospital (DH) were also engaged during Phase II of the study. Their inclusion reflected their functional role within the primary health-care ecosystem, particularly in providing referral support for severe cases, supervisory oversight, clinical mentoring, and medicine supply-chain facilitation to HWCs and PHCs. Strengthening these secondary-level units was therefore integral to reinforcing the effectiveness and sustainability of primary care delivery, rather than shifting the locus of pneumonia care away from primary care level.

A structured framework was needed to align implementation strategies with barriers, facilitators, and outcomes, to facilitate their continuous monitoring and real-time adaptation. The Implementation Research Logic Model (IRLM) serves this purpose by linking determinants to tailored strategies, clarifying assumed mechanisms of change, and defining outcome indicators for systematic evaluation.[16, 17] Logic models have been extensively used in public health to enhance the planning, execution, and assessment of implementation efforts, helping identify gaps in programme delivery and ensuring interventions are delivered with fidelity.[18] In the context of pneumonia management in India, the IRLM provides a pragmatic roadmap to strengthen primary care-led outpatient management of under-five pneumonia, while explicitly articulating the supportive roles of CHCs and the DH within referral and health-system strengthening functions.[16]

The primary aim of this manuscript is to describe co-design and iterative optimisation of an implementation model for primary care-based childhood pneumonia management in low-resource district of North India. [19] [20-22] The secondary aims are to identify to identify and map multilevel determinants-barriers and facilitators, to CPMG implementation using the CFIR framework; to describe the participatory co-design process through which context-specific implementation strategies were selected and refined using the CFIR-ERIC Barrier Busting Tool; to document the iterative development of three successive implementation models (Model 0+, Model 1, Model 2); and to present the finalised IRLM developed in collaboration with government and community stakeholders for Phase III of study. Findings from Phase III- implementation, evaluation, and district-wide scale-up, will be reported in a separate forthcoming publication.

## METHODOLOGY

This manuscript reports findings from Phase I and II of a 26-month quasi-experimental, pre-post design implementation research project aimed at developing and optimising a model for primary-care level management of under-five pneumonia in a low-performing district of North India.[23] Phase I (formative research; June-August 2023), Phase II (model development and optimization; September 2023-May 2024), and Phase III (implementation and evaluation of the optimized model followed by district-wide scale-up; May 2024-March 2025).

The current manuscript focuses primarily on Phase II, with selected findings from Phase I included only to provide context for the development of the initial implementation model (Model 0+). Detailed findings from Phase I have been reported elsewhere.[11, 15] The Phase II optimization process employed a mixed-methods, iterative design, while the quasi-experimental pre-post evaluation of the optimized model forms part of Phase III and is beyond the scope of this paper. Across the overall project, data were collected at baseline, midline, and endline, with key implementation and coverage indicators monitored fortnightly during Phase II to support real-time adaptation of the implementation model.[23] The details regarding the study surveys can be found in **Supplementary Box 1**.

### Theoretical and conceptual frameworks

The study drew on three complementary implementation science frameworks, selected following a systematic literature review of available implementation frameworks.[24]The Consolidated Framework for Implementation Research (CFIR) was selected to systematically identify and map multilevel determinants, barriers and facilitators, affecting primary care-based pneumonia management, spanning community norms, facility workflows, and health-system processes.[22] The Expert Recommendations for Implementing Change (ERIC) Barrier Busting Tool provided a structured taxonomy of implementation strategies, enabling transparent, evidence-informed, and contextually feasible strategy selection.[16, 25] The Implementation Research Logic Model (IRLM) was used to align determinants, strategies, mechanisms of change, and measurable outcomes into a coherent framework to guide programme planning, monitoring, and adaptation.[16, 20]The RE-AIM (Reach, Effectiveness, Adoption, Implementation, Maintenance) framework guided outcome specification, with explicit attention to equity and sustainability. [26]These frameworks are described in detail in **Box 1**.

#### Box 1: Description of frameworks and models used in this study

The Consolidated Framework for Implementation Research (CFIR) is a widely used determinant framework that helps identify barriers and facilitators influencing the implementation of healthcare interventions. It consists of five domains with 39 constructs that guide the systematic assessment of Intervention Characteristics: Examines aspects of the intervention, such as evidence strength and quality, complexity, adaptability, and cost, which affect its uptake.

Outer Setting: Focuses on external influences like patient needs and resources, peer pressure, external policies, and funding availability that impact implementation.

Inner Setting: Explores organizational factors such as leadership engagement, available resources, implementation climate, culture, and communication networks that facilitate or hinder implementation.

Characteristics of Individuals: Assesses the knowledge, beliefs, self-efficacy, and motivation of those involved in implementing the intervention.

Process: Evaluates how the intervention is executed through strategies like planning, engaging key stakeholders, executing, reflecting, and evaluating outcomes.

The Expert Recommendations for Implementing Change (ERIC) framework provides a taxonomy of 73 discrete implementation strategies to improve healthcare program adoption, implementation, and sustainability. These strategies include train-the-trainer models, educational outreach, financial incentives, leadership engagement, audit and feedback, and policy changes, among others.

The Implementation Research Logic Model (IRLM) is a structured framework that systematically links implementation determinants, strategies, mechanisms of change, and outcomes to guide in implementing and evaluating healthcare interventions. It ensures transparency, supports adaptation to local contexts, and facilitates evidence-based decision-making. It comprises four key components:

Determinants: Contextual factors influencing implementation, categorized under CFIR domains i.e., inner, outer, intervention, individual and process

Implementation Strategies: Targeted approaches selected using the ERIC taxonomy to address identified barriers.

Mechanisms of Change: The process by which strategies drive improvements in the primary, secondary and implementation outcomes.

Outcomes: The IRLM aligns with frameworks like Procter’s or the Reach Effectiveness Adoption Implementation Maintenance (RE-AIM) model, ensuring a comprehensive assessment of impact:

<u>Example of RE-AIM outcomes</u>: Reach: Proportion of caregivers seeking care from appropriate health facilities; Effectiveness: Adherence to pneumonia management guidelines and treatment success rates; Adoption: Proportion of healthcare facilities initiating CPMG implementation.; Implementation: Fidelity to CPMG protocols and provider adherence and Maintenance: Long-term sustainability of pneumonia management interventions.

<u>Example of Procter’s framework</u>: Implementation outcomes: Adoption, fidelity, acceptability, feasibility, and sustainability; Service outcomes: Quality, efficiency, equity, and timeliness of healthcare delivery and Patient outcomes: Care-seeking behaviour, morbidity reduction, caregiver satisfaction, and treatment adherence.

#### Box 2: Strategies identified as relevant by the research team based on the formative research findings (most of which were identified using the ERIC-CFIR barrier busting tool, except for 23 and 33 which were based on the pragmatic experiences of research team)

1. Obtaining formal commitment.
2. Centralized centre of resource procurement and distribution.
3. Network weaving among stakeholders
4. Make training dynamic
5. Involvement of high-level and mid-level leaders in active implementation of CPMG
6. Identify early adopters
7. Distribute educational materials
8. Remind healthcare providers
9. Alter incentive and allowance system for healthcare providers
10. Revise roles and responsibilities of healthcare staff
11. Conduct educational outreach camp
12. Develop and distribute IEC materials
13. Shadow other experts
14. Provide supportive supervision
15. Facilitation
16. Build coalition
17. Use of mass media
18. Conduct local consensus discussions.
19. Quality monitoring systems
20. Increase demand
21. Promote adaptability
22. Tailor strategies
23. Digitalization [Pragmatic experiences of research team]
24. Change in record system
25. Audit and feedback
26. Identify and prepare champions.
27. Ongoing training
28. Use train-the trainer strategy
29. Conduct educational meeting
30. Inform local opinion leaders
31. Capture and share information
32. Conduct local need assessment.
33. Involvement of non-RMP in implementation. [Pragmatic experiences of research team]

### Study setting

The primary implementation research (IR) project was conducted in Palwal district, Haryana, covering a population of 1,285,000 across 282 villages, 72 HWCs, 15 PHCs, six community health centres (CHCs), and one district hospital (DH). Palwal district was identified as a low-performing district in Haryana based on its suboptimal maternal and child health indicators, constrained healthcare infrastructure, and limited access to essential services, falling below state and national benchmarks for health system performance. [27, 28] Although the study area included CHCs and the DH, the implementation focus was on strengthening outpatient pneumonia management at primary-level facilities (HWCs and PHCs), with CHCs and the DH functioning as referral, supervisory, and supply-chain support units within the primary health-care ecosystem.

Phase I (time-line: 01 June to 31 August 2023) consisting of formative research including a baseline survey carried out in an area with a population of 108,000 inhabitants, including 10 HWCs, five PHCs, four CHCs, and one DH. Facilities were selected purposively by the District Health Authority based on proximity to the district hospital and availability of human resources. [11]Phase II (time-line: 15 September 2023 to 10 May 2024), focused on model optimization, conducted over seven months, within a learning cluster, a subarea covering four HWCs, three PHCs, two CHCs, one DH and a population of about 50,000. The learning cluster was selected to represent the geographic and facility-level diversity of the study area while maintaining an operational feasibility.

### Participants and sampling

Study investigators, medical doctors specialized in public health (BGP) or holding PhD (SM), conducted IDIs using semi-structured interview guides. These guides were updated regularly based on daily debriefs between the interviewers. In Phase-I and Phase-II purposive sampling was employed to select respondents such as caregivers with under-five children who had recently had signs of pneumonia, heathcare providers, and ASHAs and after obtaining verbal and/or written consent interviews were conducted. In both the phases, among eligible caregivers we selected those who were fluent in the Hindi language and/or local dialect to ensure effective communication. We invited some who sought care from government facilities, some who visited private centres, and a few who opted for home-based management. Lists of caregivers were compiled from a baseline household survey in Phase-I and from HWCs’ and ASHA records of suspected pneumonia cases in Phase-II. The purposive sampling strategy for the DHAm, healthcare providers and ASHAs differed slightly between the two phases. In Phase I, DHAm were selected based on their leadership roles in child health programs or their involvement in managing under-five illnesses at the DH. Medical Officers (MOs) from PHC were selected if they were in-charges of their respective facilities. Auxiliary Nurse Midwives (ANMs) and ASHAs were selected based on ≥5 years’ experience, local residency, and fluency in Hindi or local dialect. Since HWCs serve as the primary implementation hubs for CPMG, all Community Health Officers (CHOs), who hold leadership positions within these centres, were included in the study.[11] During Phase-II the healthcare providers and ASHAs were selected based on their performance. ASHAs’ performance was categorized based on their monthly report to ANMs of identified under-five pneumonia cases relative to the expected number of cases in the village (derived from government estimates).[8] Similarly, the performance of healthcare staff from facilities in pneumonia diagnosis was assessed as the number of diagnosed cases as per CPMG out of the total number of suspected under-five pneumonia cases visiting that centre, and the performance in management was defined as the number of cases managed as per CPMG out of the total number of diagnosed cases as per CPMG. Data for the performance assessments was obtained from the under-five outpatient department (OPD) register. For the IDIs we selected a mix of high performers (with scores ≥30% on the performance index) and low performers (with scores <30% on the performance index). The 30% threshold was selected pragmatically rather than from an established normative standard, as no validated performance benchmark existed for CPMG implementation at HWC or PHCs level at the time of study initiation. Given that the CPMG had not been implemented in Palwal prior to this study, the research team set 30% as an operational threshold reflecting the minimum expected coverage achievable within the early implementation period, based on discussion with DHAm and pilot observations during Model 0+. This threshold was used solely to ensure diverse implementation experiences were captured in qualitative sampling and was not applied as an outcome benchmark. Additionally, a few senior Medical Officers (MOs) from the CHCs were interviewed for understanding the issues pertaining to referral, supervisory, and supply-chain within these primary health-care ecosystem.

### Data collection procedures

In Phase-I and II, data collection from both caregivers and healthcare providers aimed to comprehensively assess facilitators and barriers while implementing the CPMG specifically in primary-level. The quantitative tools and qualitative guides used in Phases I and II are published elsewhere, and those exclusively used in Phase-II are attached as **Annexure 1.** of study.[11] [24] Quantitative data were obtained from four primary sources: (i) under-five OPD registers maintained by CHOs and MOs at HWCs and PHCs, recording case identification, diagnosis, management, and referral; (ii) ASHA monthly reports submitted to ANMs documenting case identification and referral activity; (iii) structured pre- and post-SBCC assessment checklists administered to caregivers; and (iv) knowledge and skills assessment questionnaires administered to healthcare providers and ASHAs.

The research team used an interview-guide to evaluate the awareness and knowledge of caregivers of under-five children of preventive strategies, pneumonia signs and symptoms, and care-seeking sources for suspected cases. The interview-guide was administered to 10% of caregivers of under-five children attending the Social and Behavioral Change Communication (SBCC) sessions, both before and after the sessions, with a seven-day interval between the pre- and post-assessments. In addition, assessments were conducted during Phase-II through quantitative questionnaires, of healthcare providers’ and ASHAs’ knowledge and skills during monthly review meetings and on-site observations. These assessments aimed to monitor the knowledge and skills gained during the initial training sessions conducted at the study’s outset and focused on critical CPMG components. Additionally, informal discussions with DHAm provided insights into systemic barriers during guideline implementation.

OPD register data quality was assured through a standardised extraction process: trained research assistants cross-checked register entries against CPMG diagnostic criteria during each fortnightly visit, flagging inconsistencies for clarification with the responsible CHO or MO. Registers were considered valid for a given fortnightly period if all required fields, date, child age, presenting symptoms, respiratory rate, temperature, diagnosis classification, treatment prescribed, and referral decision, were completed for at least 80% of recorded cases. Facilities not meeting this threshold were excluded from indicator calculations for that period and the gap was documented as a process monitoring finding. Implementation indicator assessments were standardised using a pre-specified indicator definition sheet **(Supplementary Box 2),** with all research assistants trained together at model initiation and re-calibrated at the start of each new model period to ensure inter-rater consistency in data extraction.

Sample sizes for qualitative interviews in Phase II were determined by purposive sampling to capture diversity of perspectives across cadres and performance levels, with data collection continuing until thematic saturation was reached, defined as no new CFIR-relevant themes emerging from additional interviews. The number of co-design workshops (three) was determined pragmatically based on the number of model iterations and the timeline for Phase II. To ensure rigour of the qualitative component, member-checking was employed: a summary of key emerging themes was shared with a subset of participants, two CHOs, two ANMs, two ASHAs, and two caregivers, at the end of each model period to confirm that interpretations accurately reflected their experiences. Peer debriefing was conducted through regular debriefs between the two lead investigators (BGP and SM) throughout data collection and analysis to minimise individual interpretive bias and ensure consistency of coding decisions.

### Stakeholder Engagement, co-design workshops and optimisation of the model

Three co-design workshops were planned and conducted, one at the initiation of each model iteration (Model 0+, Model 1, and Model 2), to review implementation findings and refine strategies for the subsequent model. Each workshop was preceded by a preparatory meeting in which the research team presented quantitative indicator data and qualitative findings from the preceding implementation period to DHAm, enabling evidence-informed discussion.

Upon finalising the CFIR-ERIC determinant mapping for each model period, the research team initiated formal outreach to district and state-level healthcare professionals, experts, and community stakeholders through official visits and emails, inviting them to participate in in-person co-design workshops. Stakeholders were selected based on their involvement in CPMG implementation, their technical expertise, or their role as community representatives. Key participants included Haryana state health authority members (SHAm), high and mid-level leaders from the Child Health Department and the National Health Mission, as well as DHAm including the Chief Medical Officer, Deputy Chief Medical Officer, programme officers and district-leads of the CPMG, MO in-charges of PHCs and CHCs, and the district ASHA coordinator. The final co-design workshop additionally included Panchayati Raj Institution (PRI) representatives, elected community representatives who participate directly in local decision-making, given their role in resource mobilisation, accountability, and community participation.[29]

In each workshop, Phase I and Phase II findings were presented and stakeholders were invited to identify any barriers or facilitators not previously captured. The research team then proposed implementation strategies based on the ERIC taxonomy and the CFIR-ERIC Barrier Busting Tool **(Annexure 2**), supported by practical experience. A structured discussion followed on the anticipated acceptability, feasibility, and potential challenges of each proposed strategy. Consensus was defined as agreement among most stakeholders present, with no active objection from any DHAm or SHAm representative; where dissenting views were expressed, these were documented and the strategy in question was either modified to address the concern or deferred to the next model iteration for further consideration. All workshop discussions were audio-recorded with participant consent, and a designated note-taker documented key decisions, dissenting views, and action points. Two authors with qualitative expertise (BGP and SM) subsequently coded the workshop notes and recordings using the CFIR 2.0 framework to identify any additional barriers and facilitators raised by stakeholders.[30] Where multiple ERIC strategies mapped to the same barrier, the research team presented all options to stakeholders during co-design workshops, who then ranked them by perceived feasibility and acceptability within the existing health system context; strategies receiving endorsement from both DHAm and SHAm representatives were retained, while others were deferred or dropped.

The IRLM was framed by the study team following the final co-design workshop, aligning determinants, strategies, mechanisms of change, and outcomes systematically. The team documented justifications, empirical evidence, theory-based assumptions, or practical experience, and recorded assumed mechanisms by which each strategy would influence outcomes, framed within the RE-AIM and Procter et al. implementation outcomes frameworks.[31] The IRLM was shared with stakeholders for additional inputs before being finalised and adopted for district-wide scale-up. All the findings are reported as per Standards for Reporting Implementation Studies (StaRI) checklist (Annexure 3)

### Data analysis

Quantitative and qualitative data were analysed using a mixed-methods explanatory approach to inform iterative model optimisation. Quantitative data were obtained from routine programme records (under-five OPD registers, referral slips, ASHA reports), structured checklists administered during SBCC sessions, and periodic assessments of provider knowledge and skills. Key implementation and service delivery indicators, including appropriate care-seeking, correct diagnosis, guideline-concordant management, referral compliance, and follow-up, were summarised as proportions and tracked across baseline, midline, and endline periods. Descriptive statistics were used to examine trends across successive implementation models (Model 0+, Model 1, and Model 2), primarily to guide real-time adaptations rather than estimate causal effects. Cases with incomplete OPD register entries, defined as missing data in more than 20% of required fields, were excluded from indicator calculations for that fortnightly period; the proportion of excluded records was documented as a quality monitoring metric at each assessment cycle. No imputation was applied given the programme-monitoring purpose of the analysis.

Qualitative interview data were analysed deductively using the CFIR 2.0 framework. Coding was conducted independently by two researchers (BGP and a trained research coordinator) using a pre-specified codebook developed from the 39 CFIR 2.0 constructs and adapted to the study context during a calibration exercise prior to Phase II data collection. Each transcript was coded independently by both coders. Inter-rater reliability was assessed on a random 20% subset of transcripts; discrepancies were resolved through discussion, and where consensus could not be reached, a third researcher (SM) provided a final adjudication. The codebook was updated iteratively as new contextual sub-themes emerged, with all updates documented and applied retrospectively to previously coded transcripts. NVivo 1.7.1 software was used for systematic organisation and retrieval of coded data. Qualitative findings were triangulated with quantitative indicator trends at the end of each model period to explain observed changes and inform strategy refinement for the subsequent model. All findings are reported in accordance with the Standards for Reporting Implementation Studies (StaRI) checklist (**Annexure 3**) and the Consolidated Criteria for Reporting Qualitative Research (COREQ).[32]

### Model development process and optimization

The formative research conducted during Phase I revealed that the CPMG had not been implemented in Palwal prior to the initiation of this implementation research [11] along with low community awareness of the guidelines.[15] The lack of prior implementation provided a baseline context for developing the initial implementation model, referred to as Model 0+, which was designed to address the challenges identified during the Phase-I.

Phase II aimed to explore barriers and facilitators encountered by healthcare providers primarily working at primary-level in implementing the CPMG, challenges faced by ASHAs in identifying, referring, and following up pneumonia cases, and caregivers’ experiences in recognising symptoms and seeking appropriate care following guideline initiation.

The model 0+ was optimized through a process that was recurrent, with periodic fortnightly assessments, and the **Supplementary Box 2** represents the key-indicators, recursive; by building on previous versions of the model; and iterative, progressively refining the model over eight months (**Supplementary Figure 1**: Iterative cycles of implementation model optimization). The development of the implementation strategies was done by employing the Implementation Strategy Mapping Method.[18] The steps (**Figure 1**) were: (1) In-depth interviews (IDIs) and informal discussions with healthcare providers from HWCs, PHCs, and supervisory/referral facilities (CHCs, and the DH), ASHAs and district health authority members (DHAm);(2) IDIs with caregivers of under-five children to understand challenges related to care-seeking; (3) Analysis of interview data using the Consolidated Framework for Implementation Research (CFIR) and the **Figure 2** is the illustration of implementation settings, outer context, guideline deliverers, recipients, and processes in pneumonia management in India. ;[22] (4) Utilisation of the CFIR-ERIC Barrier Busting tool to identify relevant implementation strategies,[21] (5) Co-design workshops/ meetings with DHAm and other stakeholders to review and refine proposed strategies; (6) Final revision of strategies for the optimized model based on stakeholder feedback, leading to the development of the IRLM.[33] Steps 1 to 5 were iterative and recursive, guiding the development of the optimized model from Model 0+ to Model 2 through frameworks detailed in **Box 1**. summarises the conceptual frameworks and models used in this primary care-based pneumonia management study.

**Figure 1:**
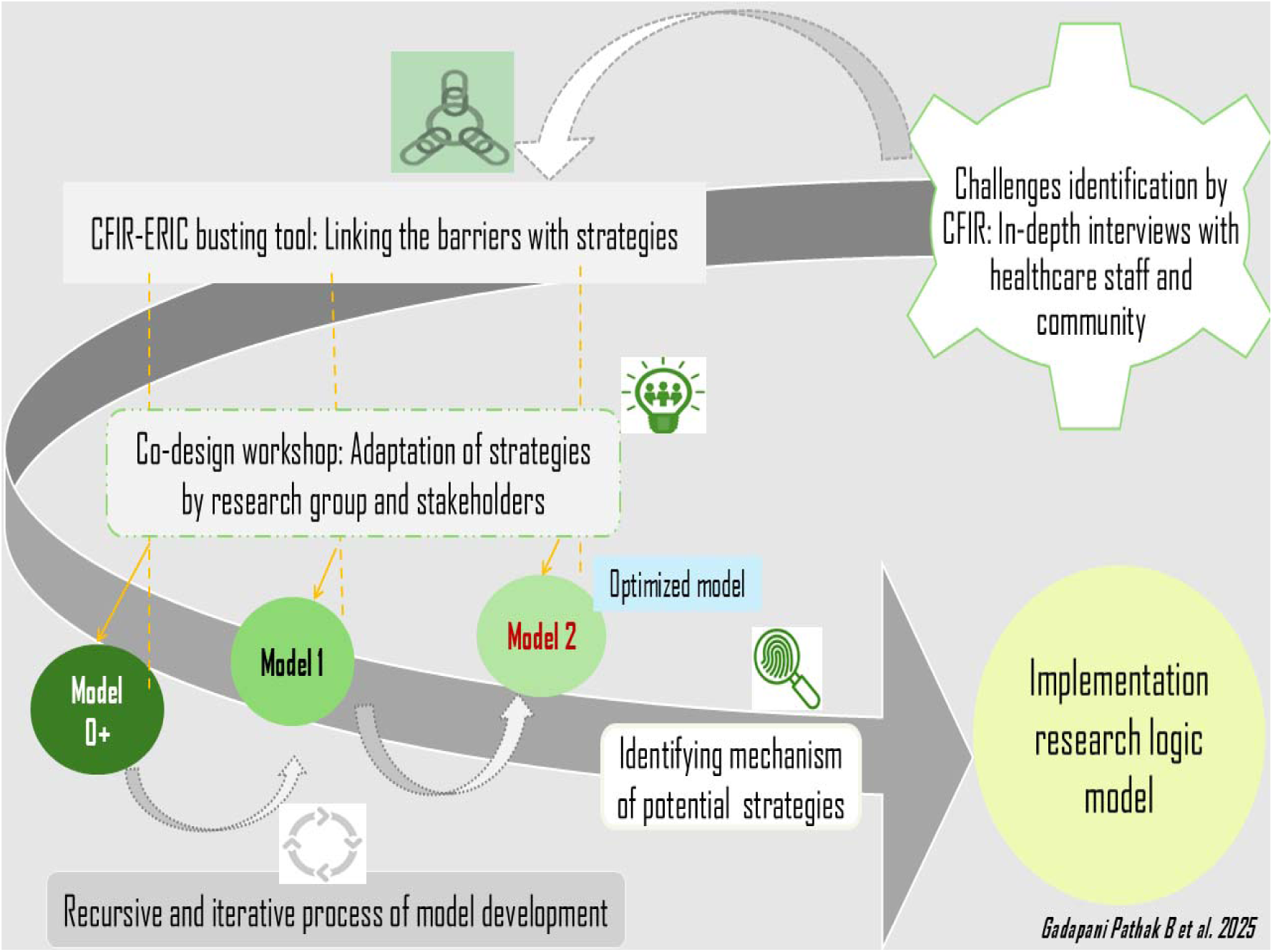
Flowchart of the development of the implementation strategy models and implementation research logic model. *CFIR: Consolidated framework of implementation research, ERIC: Expert recommendation of implementation change*.

**Figure 2:**
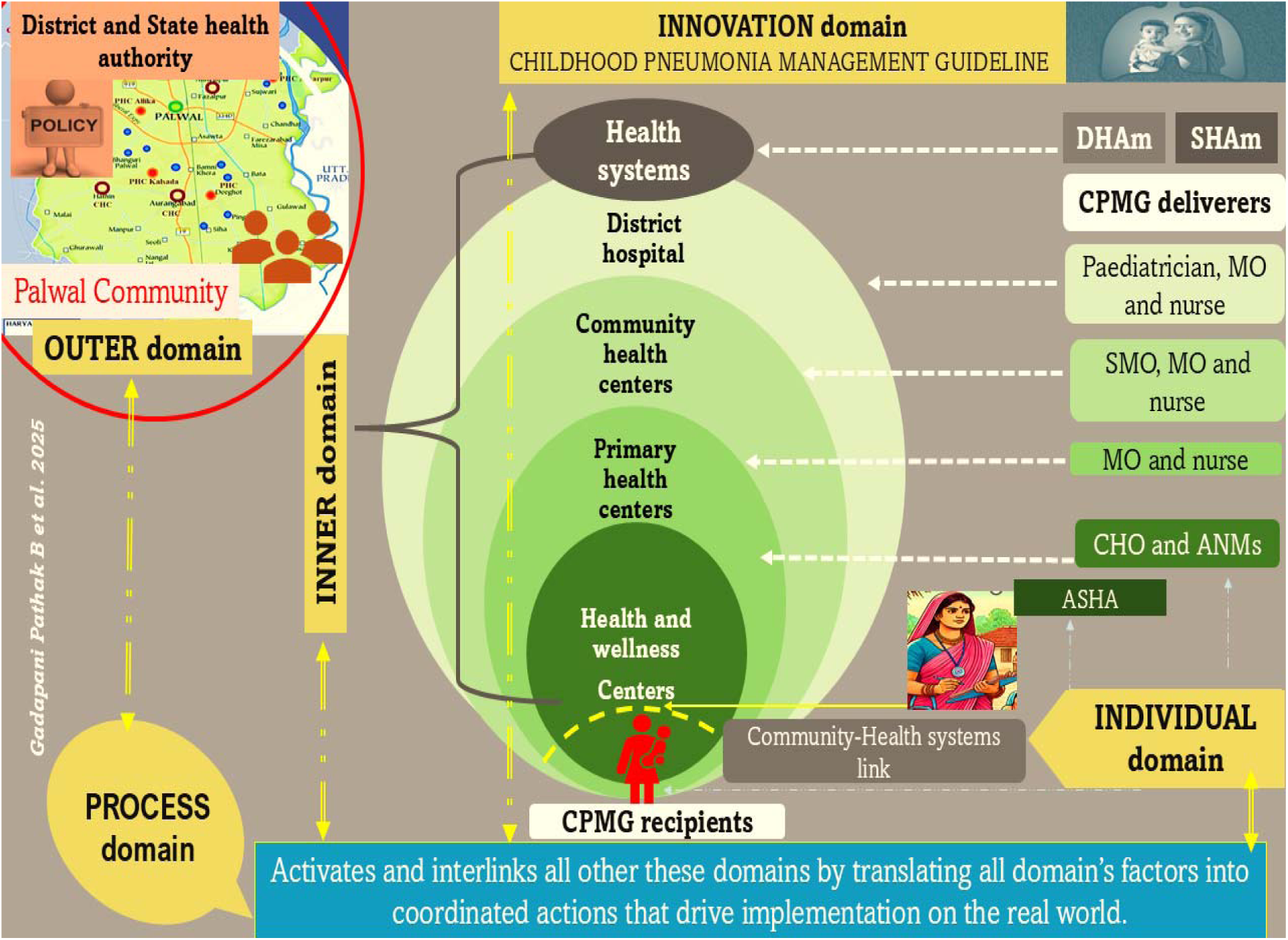
Consolidated Framework of Implementation research (CFIR)-based illustration of implementation settings, outer context, guideline deliverers, recipients, and processes in pneumonia management in India. *ANM: auxiliary nurse midwives, ASHA: Accredited social health activists, CPMG: Childhood pneumonia management guideline, SMO: Senior Medical officer, MO: Medical officer,DHAm: District health authority members, SHAm: State health authority member Childhood pneumonia management guideline logo from Palwal district map from google map image*.

We began with Model 0+, which was co-designed during a workshop in 1st week of September 2023, based on formative research findings (June-August 2023). Model 0+ was then implemented in the learning block from September 15 to November 14, 2023. Findings, including key indicator proportions and information on challenges from healthcare providers and caregivers, were documented and used to refine Model 1, which was co-designed before December 2023 and implemented from December 1, 2023, to February 28, 2024. Based on the learnings from Model 1, Model 2 was co-designed and implemented from March 8 to May 10, 2024. Model optimization was considered achieved when three key indicators surpassed 50% coverage in the learning block: (i) suspected pneumonia cases seeking care at appropriate facilities, (ii) correct diagnosis of the suspected cases as pneumonia at appropriate facilities as per CPMG, and (iii) Case management as per CPMG. This milestone marked Model 2 as the optimized version, ready for district-wide scale-up. The **Supplementary Figure-2**. represents the detailed timeline of these activities. The data-collection tools are provided in **Annexure 1** used in study Phase-II and some other published elsewhere.[11] [24]

### Implementation outcomes

The primary implementation outcome for Phase II optimisation was achievement of ≥50% coverage on all three key indicators within the learning cluster: (i) proportion of suspected pneumonia cases seeking care at appropriate primary care facilities (reach); (ii) proportion of suspected cases correctly diagnosed as pneumonia at appropriate facilities as per CPMG (effectiveness/fidelity); and (iii) proportion of diagnosed cases managed as per CPMG (fidelity to management). These indicators were selected based on the RE-AIM framework and Procter et al.’s implementation outcomes framework. Secondary implementation outcomes included: provider knowledge and skills (assessed bimonthly), caregiver awareness of pneumonia danger signs (assessed pre- and post-SBCC sessions), referral compliance, and follow-up rates. All indicators are defined in **Supplementary Box 2**. The process evaluation was conducted through fortnightly quantitative assessments of key indicators, bimonthly knowledge and skills assessments of healthcare providers and ASHAs, and pre-post evaluations of caregiver SBCC sessions, to monitor implementation fidelity and guide real-time model adaptation. No formal economic evaluation was undertaken in this phase. No sub-group analyses were pre-specified for Phase II.

### Fidelity assessment

Fidelity in this study referred to the degree to which CPMG-recommended practices were delivered as intended by healthcare providers at primary care facilities. Three fidelity indicators were defined and tracked: (i) proportion of suspected pneumonia cases correctly classified and diagnosed per CPMG diagnostic criteria by CHOs and MOs; (ii) proportion of diagnosed cases prescribed the correct antibiotic regimen i.e., oral amoxicillin for non-severe pneumonia or pre-referral gentamicin for severe cases, at the correct dose and duration; and (iii) proportion of severe cases referred to higher facilities per CPMG referral criteria. These indicators are defined operationally in **Supplementary Box 2**. Fidelity was assessed through two complementary methods: structured review of under-five OPD register entries by trained research assistants at each fortnightly visit, cross-checked against CPMG diagnostic and management criteria; and direct observation of a random sample of OPD consultations at HWCs and PHCs during monthly visits. Research assistants were trained in fidelity assessment procedures at model initiation and re-trained at the start of each new model period. A reliability check was conducted on 10% of OPD register extractions by a second independent reviewer; discrepancies were resolved through discussion and documented. Observer training for direct observation sessions used standardised checklists based on CPMG criteria (**Supplementary Box 2).**

### Relationship to companion publications

This manuscript is the third in a series of manuscripts reporting findings from the same implementation research project [14] conducted in Palwal district, Haryana. Two companion papers have previously reported findings from Phase I of the project. The first manuscript reported supply-side determinants of CPMG implementation, including infrastructure gaps, medicine supply disruptions, staffing deficits, poor record-keeping, and provider knowledge and training barriers, assessed using CFIR across facility and health-system levels.[11] The second manuscript reported demand-side determinants, including caregiver care-seeking behaviours, sociodemographic factors associated with appropriate care-seeking, and community-level barriers including reliance on informal providers, financial constraints, and gender biases.[15] Both these manuscript drew on the same Phase I dataset, participant pool, study area, and data collection instruments described in those publications. The qualitative and quantitative data collection tools used in Phase I are published in those companion papers; tools used exclusively in Phase II are provided in Annexure 1 of this manuscript. The current paper draws on Phase I findings solely as contextual background to inform the design of Phase II and presents as new results only the findings from Phase II, specifically, the barriers and facilitators that emerged during iterative model implementation (Models 0+, 1, and 2) within the learning cluster, the co-design process, and the development of the finalised IRLM. **Supplementary Table 1** provides a summary of Phase I findings from [11] and [15] for reference; the full findings are available in those publications.

## RESULTS

### Participant demographics

Phase I participant characteristics, including 53 healthcare providers, 15 caregivers, and four PRI members interviewed across the study area, are reported in full in the companion publications [11, 15] and are not reproduced here. During Phase II, 22 healthcare providers from primary care facilities (HWCs and PHCs), 15 ASHAs, and 20 caregivers of under-five children were interviewed. To understand referral pathways, admissions, and supply-chain factors influencing primary care delivery, three staff members from CHCs were also interviewed. Stakeholders participating in co-design workshops and meetings included four State Health Authority members (SHAm), ten DHAm, and three PRI representatives.

### Implementation Outcomes

The primary implementation outcomes - care-seeking at appropriate primary care facilities, correct diagnosis as per CPMG, and guideline-concordant management -improved progressively across the three model iterations within the learning cluster (**Figure 3 and Table 1).** The agreement of estimated suspected childhood pneumonia programme workload against observed facility register data detailed in **Supplementary Table 2.**

**Figure 3:**
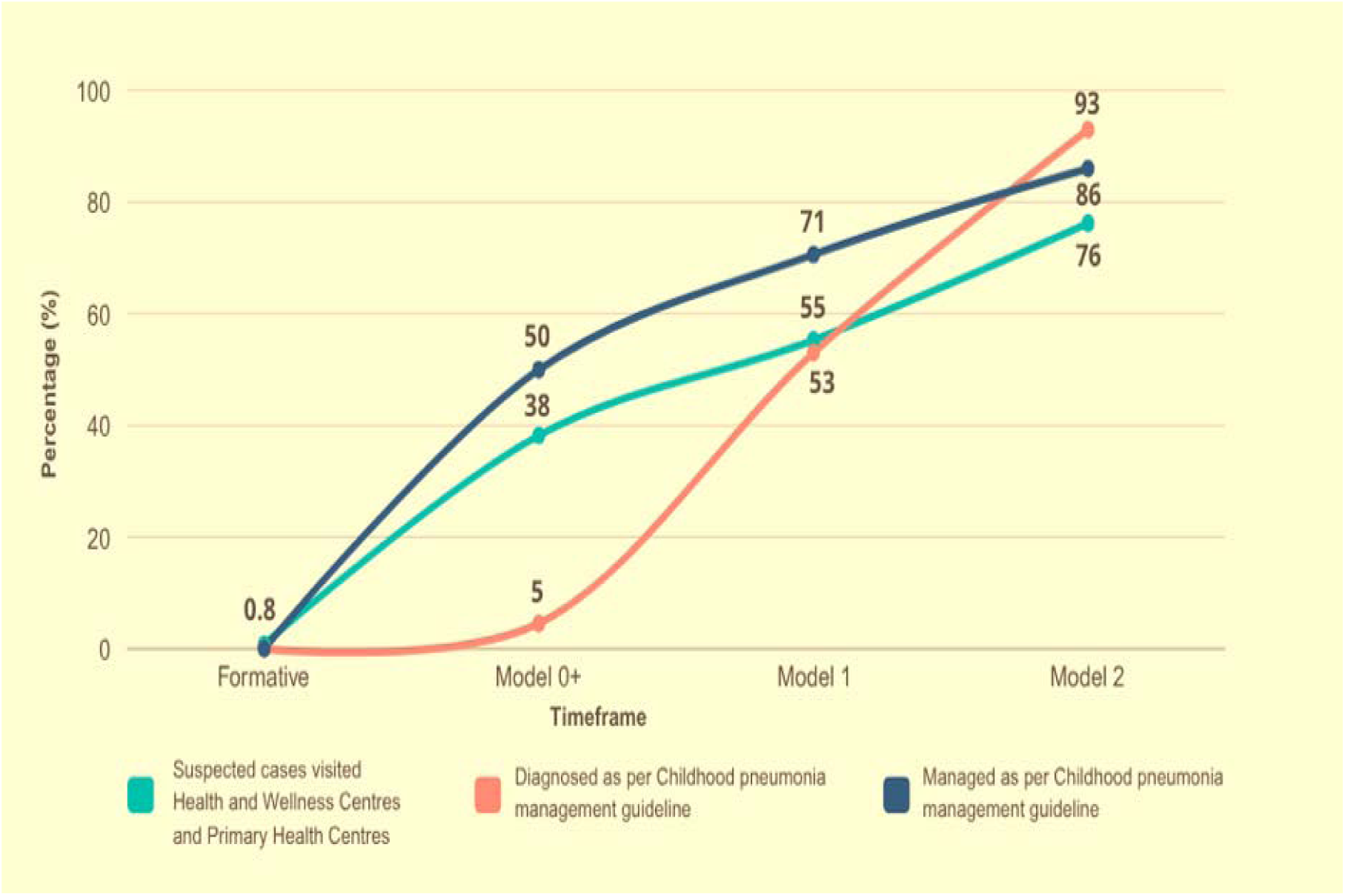
The proportion of suspected, diagnosed and managed under-five cases reaching the primary care facilities for care during Phase-I and Phase-II of study.

**Table 1:**
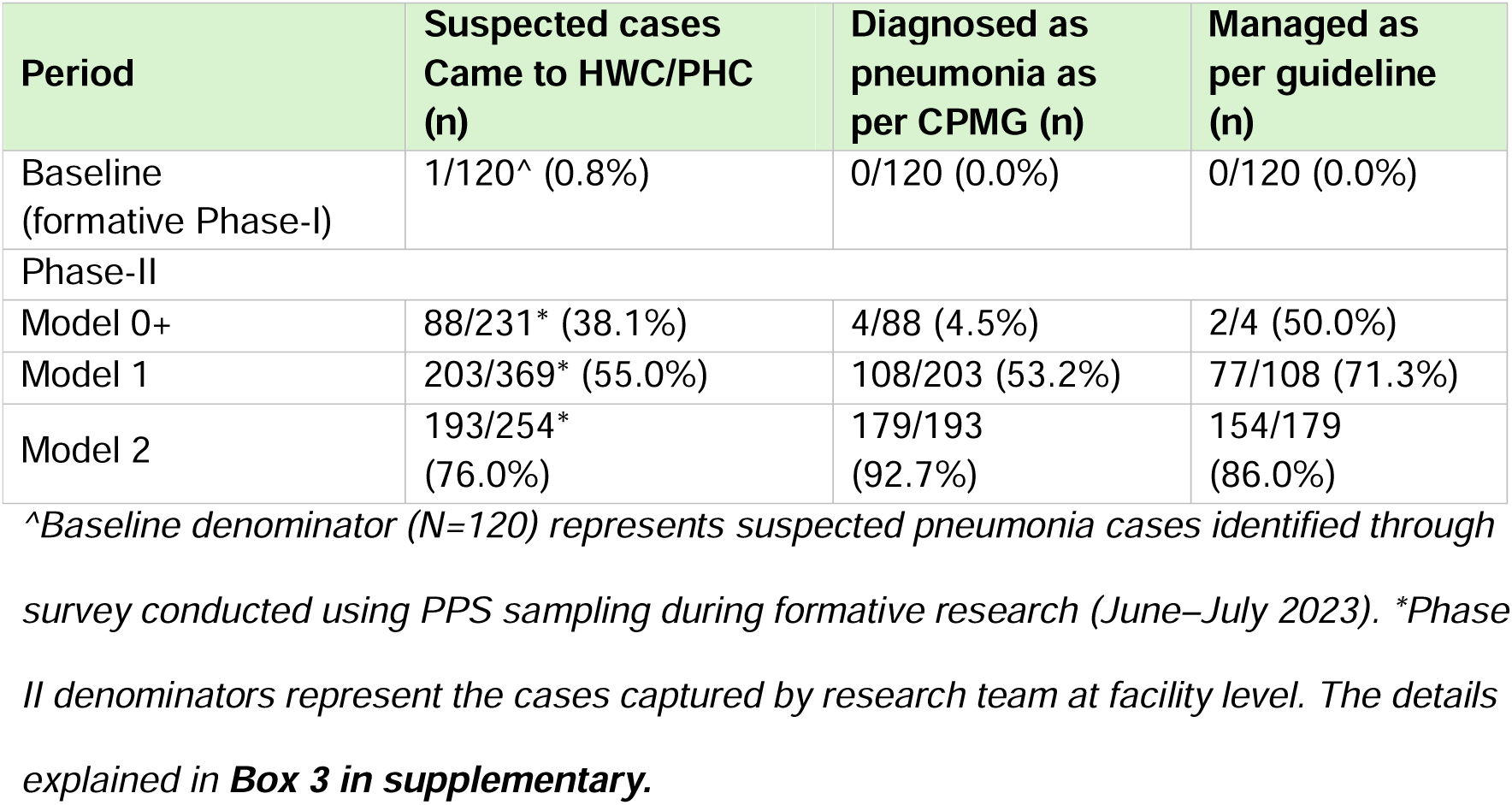
Cascade of childhood pneumonia management outcomes across implementation phases (learning cluster, Palwal district, 2023–2024)

| Period | Suspected cases Came to HWC/PHC (n) | Diagnosed as pneumonia as per CPMG (n) | Managed as per guideline (n) |
| --- | --- | --- | --- |
| Baseline (formative Phase-I) | 1/120<sup>^</sup> (0.8%) | 0/120 (0.0%) | 0/120 (0.0%) |
| Phase-II |  |  |  |
| Model 0+ | 88/231* (38.1%) | 4/88 (4.5%) | 2/4 (50.0%) |
| Model 1 | 203/369* (55.0%) | 108/203 (53.2%) | 77/108 (71.3%) |
| Model 2 | 193/254* (76.0%) | 179/193 (92.7%) | 154/179 (86.0%) |

<sup>^</sup>Baseline denominator (N=120) represents suspected pneumonia cases identified through survey conducted using PPS sampling during formative research (June–July 2023). \*Phase II denominators represent the cases captured by research team at facility level. The details explained in **Box 3 in supplementary**.

At baseline, only one of 120 suspected cases (0.8%) identified through the formative survey sought care at an HWC or PHC; no cases were diagnosed or managed as per CPMG criteria, consistent with the finding that the CPMG had not been implemented in Palwal prior to study initiation.[11] Following introduction of Model 0+, care-seeking at appropriate facilities increased to 38.1% (88/231), though correct diagnosis remained very low at 4.5% (4/88), and management fidelity was recorded in only 2 of 4 diagnosed cases (50.0%). By Model 1, care-seeking improved to 55.0% (203/369), diagnosis increased substantially to 53.2% (108/203), and management fidelity reached 71.3% (77/108). By the end of Model 2, all three primary implementation outcome indicators exceeded the pre-specified 50% optimisation threshold: care-seeking reached 76.0% (193/254), diagnosis 92.7% (179/193), and management fidelity 86.0% (154/179).

Provider knowledge scores improved progressively across all cadres from baseline to end of Phase II (**Supplementary Table 3**). The greatest improvement was observed among ASHAs, whose mean score increased from 4.6/8 (57%) at baseline to 7.4/8 (92%) by Model 2, an improvement of 2.8 points representing a 61% relative gain. CHOs improved from 5.7/8 (71%) to 7.7/8 (96%), ANMs from 5.4/8 (68%) to 6.8/8 (85%), and MOs from 6.0/8 (75%) to 8.0/8 (100%) by Model 1, maintaining perfect scores through Model 2. Notably, a slight plateauing of ANM scores between Model 0+ and Model 1 (6.1 to 6.0) was observed, consistent with the qualitative finding of fading training effects, which informed the refresher training strategy introduced in Model 2.

Among 149 caregivers assessed through SBCC pre-post evaluations across Phase II, pre-session awareness scores improved progressively from 2.9/8 (36%) at Model 0+ to 3.7/8 (46%) at Model 2, suggesting cumulative community learning beyond individual session effects (**Supplementary Table 4**). Post-session scores consistently exceeded pre-session scores at each model period, improving from 4.3/8 (54%) at Model 0+ to 4.8/8 (60%) at Model 2. No formal baseline caregiver awareness score was available prior to SBCC implementation; pre-session scores from Model 0+ therefore represent the earliest available reference point.

### Outcomes of the Implementation mapping

All findings presented in this section are original to this paper and derive from Phase II of the implementation research project. Phase I findings are summarised in the Introduction and reported in full in companion publications [11, 15]; and are not reproduced as results here.

#### a. Identifying the barriers and facilitators to implementing CPMG utilizing the CFIR

Phase I findings on barriers and facilitators to CPMG implementation have been reported in full elsewhere[11, 15] and are summarised in **Supplementary Table 1** for reference. The identified determinants served as the foundation for developing and refining implementation strategies in Phase II.

**Table 2** presents barriers and facilitators across both the study phases (I and II), mapped to CFIR construct(s) and linked ERIC strategies. The Phase I columns summarises published findings from [11] and [15], included for contextual reference; the Phase II columns present original findings from this paper. A pictorial overview of the Consolidated Framework of Implementation research (CFIR) domains and their influence on CPMG implementation across both the phases is provided in **Supplementary Figure 3**. The findings presented below are from Phase II, specifically, how barriers and facilitators evolved during iterative implementation of Models 0+, 1, and 2 within the learning cluster (September 2023-May 2024). These findings extend the Phase I evidence base by documenting how implementation challenges changed, persisted, or emerged during active model testing, and how facilitators were reinforced or newly identified during the optimisation process.

**Table 2:** Barriers and Facilitators identified across study phases using CFIR construct(s) with mapped ERIC strategies.

| CFIR domains and<br>Constructs | (Phase-I: barriers and<br>facilitators identified during<br>formative research)<br>Summarized from [11] and<br>[15] publications <sup>\$</sup> | (Phase-II: Development of optimized model) | | |
| --- | --- | --- | --- | --- |
|  | Duration: 3 months<br>(01 June - 31 August 2023) | Duration: 2 months<br>(15 September- 15<br>Nov 2023) | Duration: 3 months<br>(01 December-28<br>February 2024) | Duration: 2 months<br>(08 March-10 May 2024) |
| DOMAIN: INNER |  |  |  |  |
| **Construct:** Structural Characteristics |  |  |  |  |
| i. Physical Infrastructure<br>1,5,11,20,29 | - Some HWCs operate<br>within private premises,<br>making it challenging for<br>caregivers to identify the<br>centers. |  |  |  |
|  | -Small examination rooms, absence of proper examination tables and chairs for patients and caregivers<br>-A few HWCs had privacy issues e.g. male under-five caregivers had to wait outside when antenatal women were being examined to avoid compromising privacy. |  |  |  |
| ii. Work Infrastructure<br>4,5,6,10,22 | - A subset of CHOs reported instances of competing activities on the same day, with both ANMs and CHOs having tasks | - The ASHAs were finding the awareness generation activities, case-identification | -ASHAs felt overwhelmed conducting SBCC activities for pneumonia on | -MOs at PHCs and CHCs were unable to complete the under-five registers due to the high volume of patients. |
|  | elsewhere, resulting in the occasional temporary closure of HWCs.<br>-Most ANMs believed that under-five examinations and management were primarily the responsibility of CHOs, not theirs<br>-Most of the ASHAs reported feeling overwhelmed by their involvement in multiple national programs and viewed suspected pneumonia case identification and referral as an added burden | and follow-up overwhelming. | designated days, as they were already engaged in multiple awareness initiatives through platforms such as Village Health and Nutrition Days etc. |  |
| iii. Information Technology infrastructure<sup>22</sup> | Most healthcare workers indicated that they did not use digital technology for following up diagnosed pneumonia cases. |  |  |  |
| **Construct:** Communication<sup>3,5,15,22</sup> |  |  |  |  |
|  | Lack of official communication about non-functional equipment, intermittent drug supplies and other issues in all levels of healthcare. | ASHAs encountered challenges in formally communicating guideline implementation issues to the district ASHA coordinators. |  |  |
| **Construct:** Available resources |  |  |  |  |
| i. Material and equipment<br>1,2,3,4,19 | -Interrupted medicine supplies to most of the HWCs and PHCs.<br>-A few centers had non-functional equipment like pulse-oximeters, and other essential items: thermometers and weighing scales.<br>-Non-availability of IEC materials displaying information on under-five pneumonia in most of the centers. | - HWCs primarily depended on CHCs for medicine supply. However, with multiple HWCs under each CHCs, medicine requests often went unfulfilled due to overburdening of the system and mismanagement of requests.<br>- A few of the HWCs lacked supply of nebulizers. | -The study found that refresher training on pneumonia diagnosis and management was crucial, as CHOs and ANMs showed declining scores in knowledge assessments conducted every four months by the research team. However, the sustainability of these trainings was challenged by the absence of a | - Although untied funds and budget reallocation ensured a steady medicine supply across all healthcare levels, the research team observed supply disruptions again during April-May.<br>-Some CHOs were observed administering antibiotics without a confirmed pneumonia diagnosis. This practice stemmed from the approaching expiry dates of antibiotics and persistent pressure from certain caregivers to receive the |
|  |  |  | dedicated skill-station in district hospitals for continuous capacity building. | medication |
| ii. Funding <sup>1,7,11,12,14,27</sup> | -The district health authority members reported that the CPMG budget had funds for organization of trainings, printing of training modules and guidelines, IEC material development, SBCC activities, logistics | -Funds allocated within the district CPMG budget had designated heads for logistics and supplies, including items such as nebulizers. (Facilitator) | -Some district health authority members noted that more funds were allocated for IEC activities than what was usually necessary, as materials like banners did not require annual | -Funds were found to be available in the district level budget for skill station establishment (Facilitator) |
| | and supplies like pulse oximeters and monitoring and evaluation activities. (Facilitator)<br>-Availability of untied funds (583 \$/annum) for each HWCs for procurement of equipment, infrastructure upgradation, and medicine purchase. (Facilitator) | | replacement in healthcare facilities. The officials mentioned that this provided an opportunity to reallocate these surplus funds to other CPMG-related activities. (Facilitator) | |
| **Construct:** Access to knowledge and information<sup>7,8</sup> |  |  |  |  |
|  | -There was a lack of clear guidance and limited access to guideline materials essential for enhancing knowledge and | -The CHOs and ANMs faced challenges in retaining the recommended | Outdated referral slips with the ASHAs. |  |
|  | ensuring the appropriate delivery of interventions as per the CPMG among the ANMs, CHOs and MOs. | dosage guidelines outlined in the CPMG for appropriate management.<br>-No referral slips in the healthcare facilities.<br>-No job aids for counselling the caregivers. |  |  |
| **Construct:** Relative priority<sup>1,5,6,29</sup> |  |  |  |  |
|  | -Facilities at all levels lacked a sense of urgency or priority in implementing the CPMG, as it was not emphasized at the district or state levels. |  |  |  |
|  | -Many MOs recognized the importance of appropriate pneumonia management and expressed that the CPMG could play a significant role in improving child health outcomes.<br>(Facilitator) |  |  |  |
| **Construct:** Culture |  |  |  |  |
| Learning-centeredness<sup>13,14,15</sup> | At several PHCs and among district health authority members, the research team observed a proactive attitude toward improving practices and adopting a data-driven |  | -At the HWCs the CHOs were found to calculate the amoxicillin and gentamicin dosage required for treating the estimated number |  |
|  | approach to clinical decision-making.<br>(Facilitator) |  | of cases and to submit a drug procurement request at least 1 week prior before month ending<br>(Facilitator) |  |
| Recipient-Centeredness<sup>1</sup>,<br>5,9,10 | Lack of recipient-centred approach among ASHAs and ANMs, as they were involved in the implementation of multiple programs. A few examples were: at HWCs, there was no designated paediatric space; children were examined in open areas or next to antenatal women by the ANMs making caregivers uncomfortable and hesitant. ASHAs were observed to provide rushed or incomplete counselling on signs and symptoms of pneumonia and were unable to answer caregiver questions meaningfully. |  |  |  |
| Human equality centredness. <sup>11, 12</sup> | Most centers, healthcare staff and ASHAs demonstrated shared values and norms emphasizing equal treatment for all individuals, regardless of gender, race, religion, or socio-economic status. (Facilitator) |  |  |  |
| DOMAIN: OUTER SETTING |  |  |  |  |
| Construct: Critical incidents <sup>1,5,11,17,20</sup> |  |  |  |  |
|  | -Most District health authority members mentioned that the implementation of CPMG in the state of Haryana was hindered primarily due to the Covid-19 pandemic.<br>-Many caregivers refrained from visiting government centers for child illnesses, fearing that the substantial influx of people with Covid-19 in government hospitals would increase the risk of<br>their children falling sicker. |  | -The central medicine depot in Faridabad faced a shortage of antibiotics, potentially linked to disruptions caused by preparations for the ongoing state elections. The supply to healthcare centers was not completely halted, but there were noticeable delays in the delivery timeline. | -The retirement of the Chief Medical Officer of Palwal district, a key implementation leader for CPMG, and the subsequent delay in filling the position for several months was observed to be a disruption the smooth implementation of the CPMG.<br>-Ongoing shortage of medicines due to preparation of the state elections. |
| **Construct:** Local attitude<sup>11,12,16,17,34</sup> |  |  |  |  |
|  | -Strong beliefs in faith-healer or non-RMP for curing under-five pneumonia.<br>-Stigma of using services like ambulance for transportation of cases of pneumonia<br>-Preference for private care facilities.<br>-Delay of treatment because pneumonia considered non-severe childhood disease<br>-Non-compliance to | -Some caregivers insisted on antibiotics for their child, even in the absence of a pneumonia diagnosis.<br>-The research team observed that caregivers specifically from better socio-economic status still preferred private facilities over | -Some caregivers were observed administering medicines to their children for only three days instead of the prescribed five.<br>Further interviews with families revealed that some HWCs had provided only a three-day supply of medication. | -Caregivers were observed purchasing medicines from private pharmacies, and they expressed frustration and demotivation as they had to incur out-of-pocket expenses for drugs that were supposed to be freely available at government health facilities.<br>Additionally, some caregivers sought care from private centers, primarily due to incomplete medicine dosages provided at HWCs |
|  | treatment duration due to caregivers' beliefs that medicines are required only until symptoms disappear.<br>-Delay in seeking care for girl children, specifically households with multiple girl children. | government ones due to their flexible operating hours and shorter waiting times.<br>-A few well-regarded private facilities were commonly chosen as a second choice of care, particularly when the management provided by government facilities did not yield the desired improvement in the child's<br>condition. |  |  |
| **Construct:** External Pressure |  |  |  |  |
| Performance pressure <sup>1,19</sup> | -The research team noted the absence of performance pressure, such as audits or mandatory reporting of pneumonia cases, for providers of different facilities by the district health authorities.<br>-District health authority members mentioned that the CPMG outlines specific performance indicators for monitoring performance. | The research team observed that some centers experienced lower footfalls of suspected cases, and the under-five outpatient department registers were poorly maintained. Despite this, no negative feedback was given by the district health authorities regarding the performance of |  |  |
|  | (Facilitator) | these HWCs. |  |  |
| Societal pressure<sup>17,20</sup> | Due to limited community awareness about under-five pneumonia management, there was no observed experience of societal pressure on healthcare providers from the community. |  | Several CHOs reported an increase in caregiver footfall at HWCs, with more caregivers seeking treatment for suspected under-five pneumonia.<br>(Facilitator) |  |
| DOMAIN: INDIVIDUAL |  |  |  |  |
| Construct: Need<sup>1,5,6,29</sup> | -A few senior MOs stressed on opting for an integrated approach to address childhood diseases rather than<br>relying on a single program for a specific disease.<br>-Most of the CHOs felt the need of implementing child health programs, highlighting that enhancing the capacity of HWCs to deliver quality care, including the provision of medicines and consultations closer to caregivers' homes, would benefit all families, particularly the underprivileged, and address the preference for private facilities. |  |  |  |
|  | (Facilitator)<br>-Most of the caregivers felt the need to obtain a consultation and appropriate management for under-five children near to their home (Facilitator) |  |  |  |
| Construct: Capability<sup>6,7,28</sup> | - There was a lack of knowledge regarding the signs and symptoms of under-five pneumonia, insufficient skills for accurately diagnosing pneumonia cases in children, challenges in calculating the appropriate dosage based on the<br>child's age, and limited understanding of referral criteria as outlined in the guideline<br>recommendations among most of the healthcare staff.<br>-Limited awareness among the under-five caregivers about signs of pneumonia and the services available for children in the HWCs. | The CHOs were not confident to administer gentamicin.<br>- The community was having little knowledge on the preventive aspects of pneumonia. |  | The community was still having little knowledge on the danger signs of pneumonia. |
| **Construct:**<br>**Opportunity**<sup>4,7,14,29,11,12,17,20</sup> | -Most of healthcare staff had not received formal training on the CPMG.<br>Among those trained, like a few ASHAs, CHOs and | -The IEC materials lacked preventive information on under-five pneumonia and the community's |  |  |
|  | MOs, there was a notable lack of hands-on, practical training to reinforce their understanding and skills.<br>-Most caregivers of under-five children reported not receiving any awareness sessions or counselling on the signs and symptoms of pneumonia. | awareness were limited on this aspect. |  |  |
| **Construct:** Motivation<br>9 15,20,26 , | - Most of the ASHAs were not motivated to implement the guideline specifically due to the lack of incentives for case identification, referral and follow-up of cases. | -The research team observed a shift in care-seeking behaviour, with HWCs emerging as the preferred choice for suspected | -Empowered caregivers who visited HWCs for free consultations and treatment for their suspected under-five pneumonia children |  |
|  | -The ANMs also expressed feeling overburdened with the implementation of numerous other national programs and maintaining records for each of them. Hence, they were not motivated to take on an additional responsibility for a new childhood program.<br>-The CHOs, and MOs were found to be motivated to implement the CPMG as they desired to provide quality care to the community and most of them did not expect any<br>incentives/rewards<br>(Facilitator)<br>-A few CHOs experienced mistreatments from the community, which demotivated them. The caregivers had been upset due to unavailable medication or the need for referral due to stockouts. | pneumonia cases. Most of the caregivers reported that the availability of proper consultations and free medicines motivated them to seek care at these facilities. (Facilitator) | expressed dissatisfaction, as some reported receiving only a three-day supply of medication instead of the full five-day course. |  |
| Domain: PROCESS |  |  |  |  |
| Teaming <sup>22</sup> |  | A lack of coordination and collaboration in executing interdependent tasks was observed among healthcare staff,<br>particularly between ANMs and CHOs, as well as between MOs and staff nurses, and this hindered smooth implementation of the national program's activities. |  |  |
| Planning<sup>21,22, 23</sup> |  | There was an absence of clearly defined steps to address challenges in most of the PHCs such as staff being overburdened with patients, inability to fulfil the roles and responsibilities outlined in the CPMG with fidelity, and the lack of referral linkages of severe cases. |  |  |
| Reflecting and evaluation<sup>1,24,25</sup> |  | -The healthcare providers were motivated to use a data-driven approach for intervention delivery. (Facilitator)<br>- However, most centers lacked systematic data collection on specific pneumonia indicators<br>and did not utilize available data to strengthen the implementation of any components of the CPMG interventions. |  |  |
| Assessing context<sup>26</sup> | A few MOs assessed community needs during their field-visits, focusing on childhood illnesses (Facilitator). |  | A few ASHAs reported that in certain low-literacy pockets, caregivers struggled to understand the signs, symptoms, and preventive strategies for pneumonia. To address this, ASHAs began using pictorial pamphlets to communicate key messages. They observed an improvement in understanding among these communities following this strategy. (Facilitator). |  |
| Domain: INNOVATION (CPMG) |  |  |  |  |
| Source<sup>1,5</sup> | Since the CPMG was introduced by the Indian government, most healthcare providers felt obligated to implement the guidelines and fulfil their associated responsibilities<br>(Facilitator) |  |  |  |
| Complexity<sup>4,22,27,34</sup> | -Most of the CHOs found administration of the pre-referral gentamicin dose to children under five to be a complex procedure.<br>-ANMs expected that they would encounter difficulties in tracking and administering gentamicin to non-compliant children who did not visit HWCs to complete the recommended five days of injection gentamicin. | ANMs faced challenges in administering gentamicin, particularly on holidays like Sundays, for cases where caregivers refused referral to higher centers. | -Ensuring proper case management amidst interruptions in the supply of medicines was complex for most of the CHOs. |  |
| Cost<sup>1</sup> | The district health authority members reported that<br>there was no specific<br>budget head for<br>the procurement of<br>antibiotics in the CPMG. |  |  |  |
| *<sup>\$</sup>Phase I findings are reported in full in references [11] and [15]. This column is reproduced here in summary form only, to provide context for the Phase II strategy development reported in this paper.* | | | | |
| *The number marked against the barriers and facilitators are the proposed implementation strategies as per the ERIC recommendations and research team's pragmatic experiences elaborated in Box 2.* |  |  |  |  |

##### a.i. Inner setting domain

During Model 0+ implementation, several inner setting challenges persisted or newly emerged within the learning cluster. HWCs remained heavily dependent on CHCs for medicine supply, and the high HWC-to-CHC ratio led to mismanagement of requisition requests and intermittent stock gaps that had not been fully resolved by the time of model initiation. A few HWCs also lacked nebulizers, which were not covered by initial procurement. ASHAs reported that pneumonia awareness, case identification, follow-up, and SBCC activities were assigned as standalone tasks rather than being integrated into existing routine platforms such as Village Health and Nutrition Days, creating additional workload burden. Referral slips and counselling job aids remained unavailable for ASHAs and CHOs at model initiation, and high patient loads limited MOs’ ability to maintain dedicated under-five OPD registers.

A new challenge that emerged specifically during Phase II was the fading of training effects over time. Post-training knowledge assessments conducted three to four months after the initial CPMG training revealed declining recall of pneumonia classification criteria and management protocols among CHOs and ANMs, a finding not assessable during Phase I, which preceded model implementation. During Models 1 and 2, the research team observed that some CHOs prescribed antibiotics without a confirmed pneumonia diagnosis, citing concerns about approaching drug expiry dates and persistent caregiver pressure to receive medication; this pattern of antibiotic prescribing outside guideline criteria emerged as a Phase II-specific implementation fidelity concern requiring targeted strategy adaptation. A dedicated clinical skill station recommended under the CPMG for ongoing provider mentoring had not been established at the district hospital by February 2024, limiting the capacity for continuous clinical supervision of primary care providers throughout the optimisation period.

Key enabling factors observed during Phase II included the availability of designated CPMG budget funds for logistics procurement during Model 0+, and the reallocation of surplus IEC funds during Model 1 to procure essential equipment including nebulizers. These represented adaptive resource management responses that were facilitated by the formal commitment obtained from DHAm during co-design workshops.

##### a.ii. Outer setting domain

During Model 0+ implementation, community attitudes continued to shape care-seeking in ways that created new implementation challenges beyond those documented in Phase I. Some caregivers actively requested antibiotics from CHOs and MOs even when no pneumonia diagnosis had been made, creating pressure on providers to deviate from guideline-concordant management. This pattern was not documented in Phase I because the CPMG had not yet been implemented at that time.

A new and previously unobserved challenge emerged during Model 1: a few caregivers administered prescribed antibiotics to their children for only three days rather than the guideline-recommended five days. On further inquiry, the research team found that some HWCs had dispensed only a three-day supply of medication due to insufficient stock, a medicine supply problem that manifested differently from the baseline stockouts documented,[11] because it now occurred within an active implementation context where demand had increased.

The research team also observed that, even after two months of Model 0+ implementation, DHAm had not provided corrective feedback to facilities with low suspected pneumonia case footfall or poorly maintained OPD registers, indicating that performance monitoring and accountability mechanisms remained weak despite the formal commitment strategies implemented at model initiation.

During Model 2, new contextual disruptions affected the outer setting. Notably, the availability of untied funds ($575 annually per primary-level centre) and budget reallocation of surplus IEC funds had initially stabilised medicine availability during Models 0+ and 1, representing a significant enabling factor in sustaining implementation momentum. The retirement of the Chief Medical Officer of Palwal, a key implementation leader, and a months-long delay in appointing a replacement coincided with state election preparations, temporarily weakening district-level leadership engagement. Concurrently, the central medicine depot faced antibiotic shortages potentially related to election period disruptions, causing supply interruptions that resurfaced despite earlier stabilisation during Models 0+ and 1.

##### a.iii. Individual domain

During Model 1 implementation, CHOs continued to report a lack of confidence in administering injection gentamicin independently, particularly for pre-referral doses. This persisted despite initial training and reflected an ongoing need for supervised practice that the initial training alone had not addressed. Most ASHAs remained demotivated by the absence of financial incentives for pneumonia case identification, referral, and follow-up-a barrier first identified in Phase I [11] that had not been resolved by Model 1 strategies, and which the research team flagged for more targeted strategy adaptation in Model 2.

Caregivers’ awareness of pneumonia danger signs, assessed through qualitative interviews at the end of Model 1 (approximately four months into CPMG implementation), remained low, and the research team observed that prevention of pneumonia was being inadequately covered by ASHAs during SBCC sessions, suggesting that the content and quality of community education activities required refinement beyond what had been initially implemented.

A notable positive shift was observed during Model 2. By the seventh month of CPMG implementation, the under-five OPD registers at HWCs in the learning cluster reflected a meaningful increase in caregivers seeking care for suspected pneumonia at government primary care facilities. Qualitative interviews with caregivers confirmed that access to free consultations and medicines at HWCs had become a motivating factor for this shift, a change in community behaviour that had not been observed in Phase I when no model was in place.[15] However, caregivers from higher socioeconomic backgrounds continued to prefer private facilities due to flexible operating hours and shorter waiting times, indicating that the shift in care-seeking was not uniform across socioeconomic groups.

##### a.iv. Process Domain

Challenges in team coordination persisted during Phase-II of the study, particularly between ANMs, CHOs, and MOs, particularly in facilities where role boundaries remained unclear, hindering smooth implementation of primary care-based pneumonia services. The absence of clearly defined protocols for managing competing workload demands, fulfilling CPMG-specific responsibilities with fidelity, and maintaining referral linkages for severe cases remained a concern throughout Models 0+ and 1, requiring iterative strategy refinement-. During model 0+ implementation, most facilities lacked systematic data collection on pneumonia-specific indicators e.g. number of pneumonia cases identified, managed, referred, or followed-up, but the healthcare provider

s expressed willingness to use such data for performance monitoring, which was a facilitator that supported the introduction of dedicated OPD registers and audit-and-feedback mechanisms in subsequent models.

##### a.v. Innovation domain

Managing pneumonia cases amid medicine supply interruptions remained complex for CHOs throughout Phase II. ANMs struggled with tracking and administering gentamicin to non-compliant cases refusing referrals, particularly on Sundays when HWCs were closed. However, since documented instances of denied referrals, refusals were few (negligible), this challenge did not substantially affect the overall fidelity to guideline-based management at the learning cluster level.

#### b. Linking the CFIR barriers and facilitators with ERIC strategies

A total of 33 implementation strategies were identified as contextually relevant using the CFIR-ERIC Barrier Busting Tool, 31 from the original ERIC taxonomy and two additional ones derived from literature and the study team’s pragmatic assessment of feasibility and local relevance. These strategies were theoretically linked to identified barriers and facilitators. Some were one domain-specific, while others influenced multiple CFIR domains and were prioritized during co-design for their potential to strengthen CPMG implementation.

One key strategy was obtaining formal commitment from leadership, expected to influence several domains: inner setting (e.g., access to resources), outer setting (e.g., policy alignment), individual (e.g., staff motivation), and process (e.g., performance monitoring). Another widely adopted strategy was the distribution of educational materials, including guideline summaries, IEC charts, and facilitator booklets in local languages. This aimed to address knowledge-related barriers across inner, outer, and individual domains by supporting guideline adherence among CHOs and ANMs and improving caregiver understanding of pneumonia symptoms and care pathways. While these assumptions were grounded in stakeholder feedback, they were not formally evaluated.

Revision of patient record systems was also prioritized to address gaps in data quality and performance tracking at primary care facilities. At baseline, under-five pneumonia cases were inconsistently recorded, limiting the ability to monitor trends, estimate medicine needs, or assess provider performance. The introduction of dedicated pneumonia registers and standardized referral slips was expected to improve documentation and support routine quality monitoring.

#### c. Co-design review meetings to discuss the implementation strategies

During three co-design workshops held in Phase II, DHAm, SHAm, and community representatives reviewed the proposed implementation strategies and collaboratively identified those deemed most feasible and acceptable within the existing health system context. These were then operationalized (**Table 3**: Co-designed strategies with the stakeholders during the district and state review meetings and workshops). Many strategies were adapted over time, some showing early success, while others required iterative refinement across Models 0+, 1, and 2. The final strategy bundle, integrated into Model 2, represented the optimized version for strengthening primary care-based pneumonia management. This participatory, iterative refinement process helped ensure better alignment with real-world conditions and stakeholder expectations, as reflected in the Implementation Research Logic Model (**Table 4** which represents implementation research logic model for achieving high coverage of appropriate management among under-five children suffering from pneumonia).

**Table 3:**
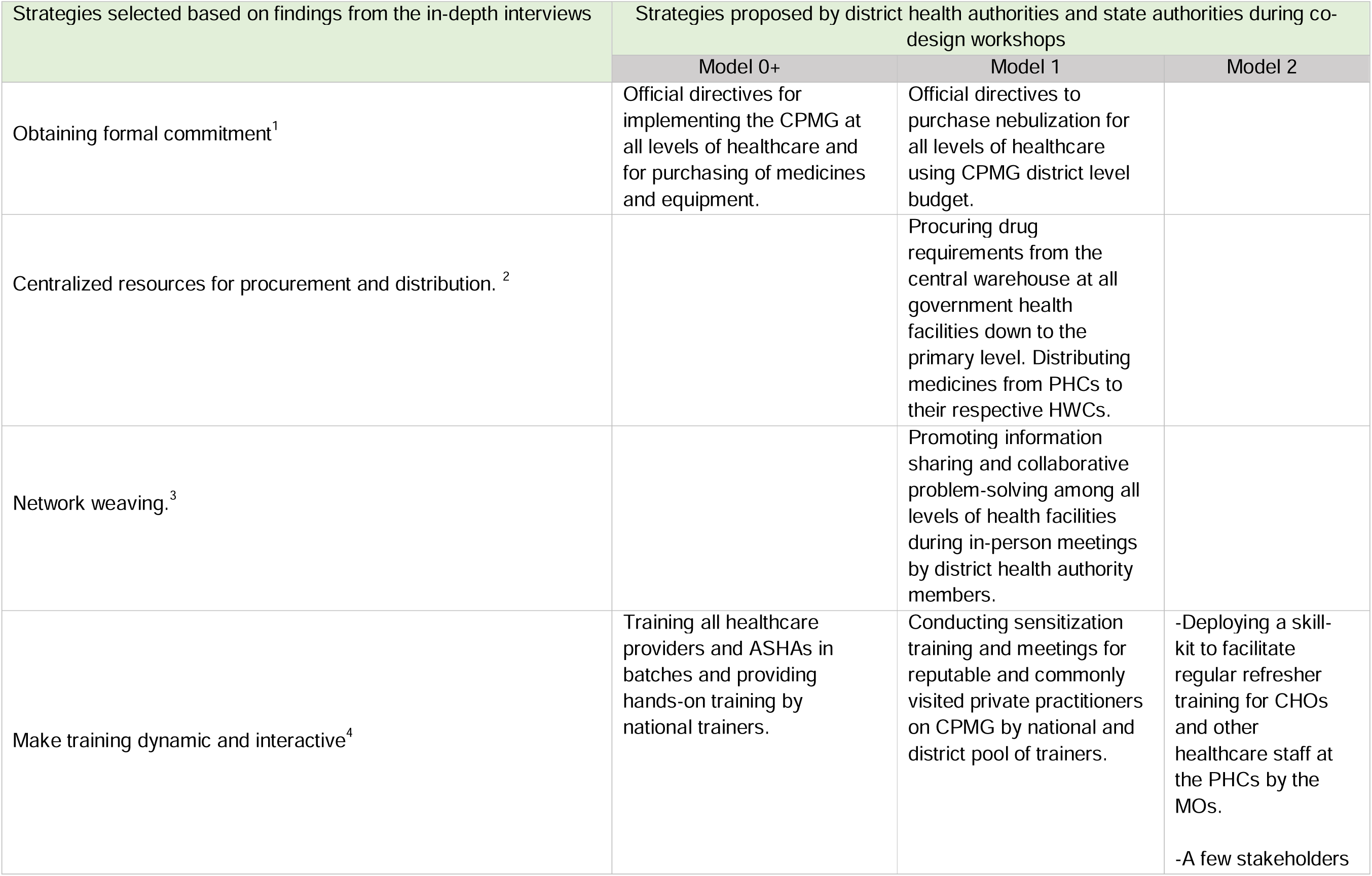
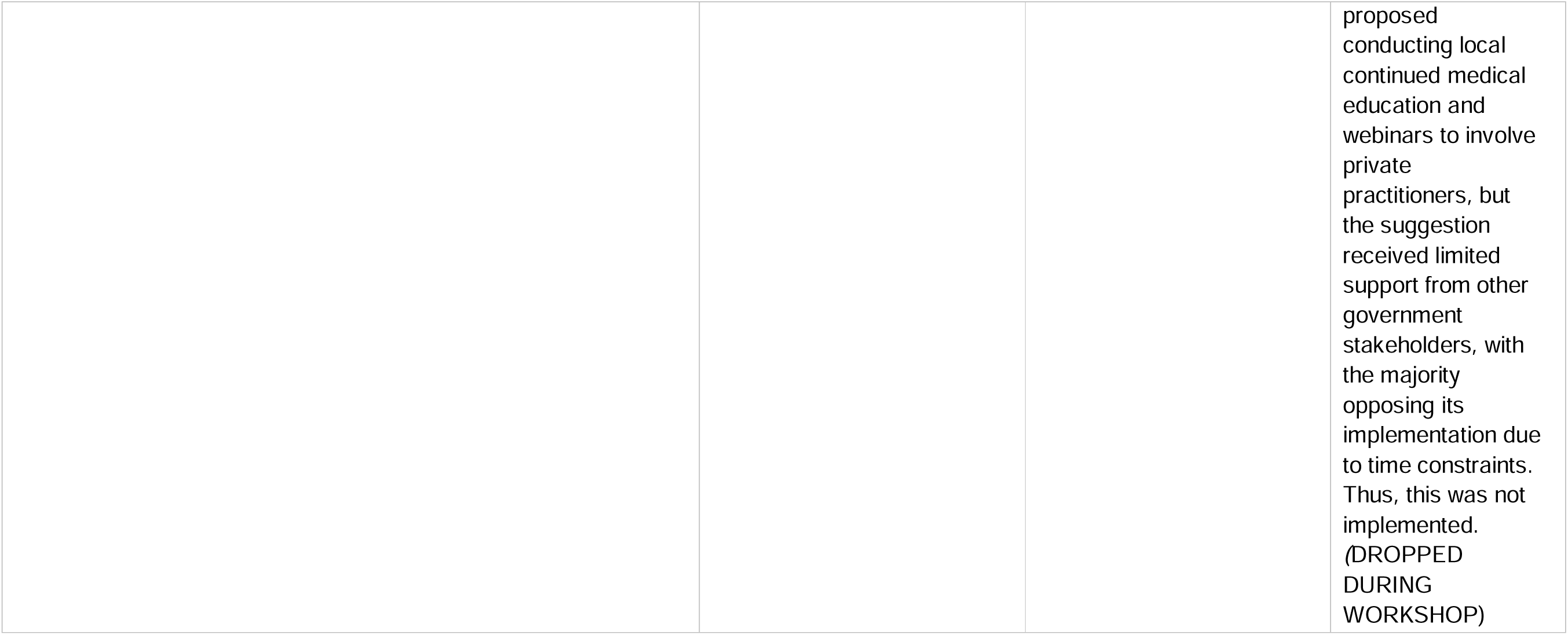
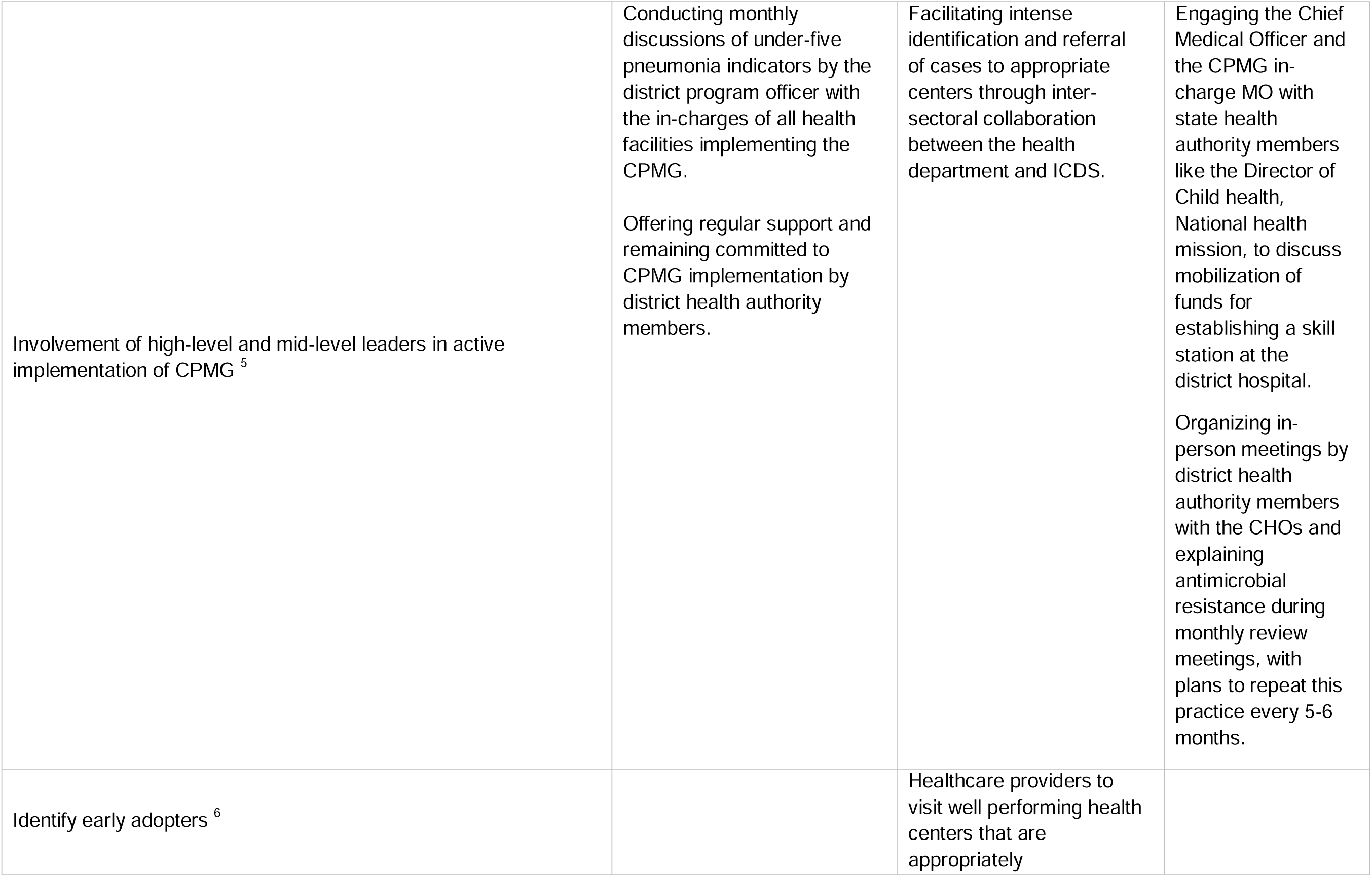
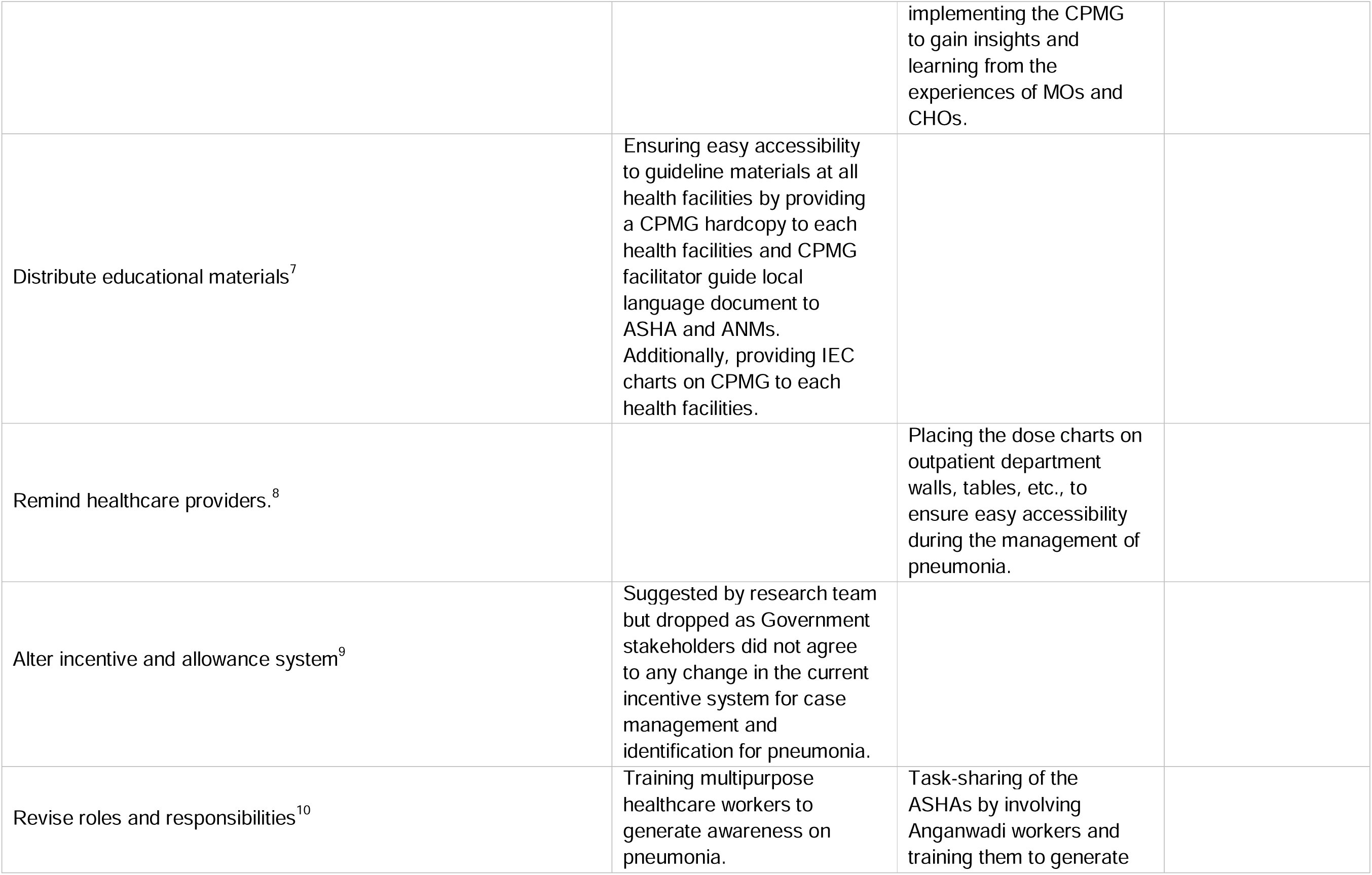
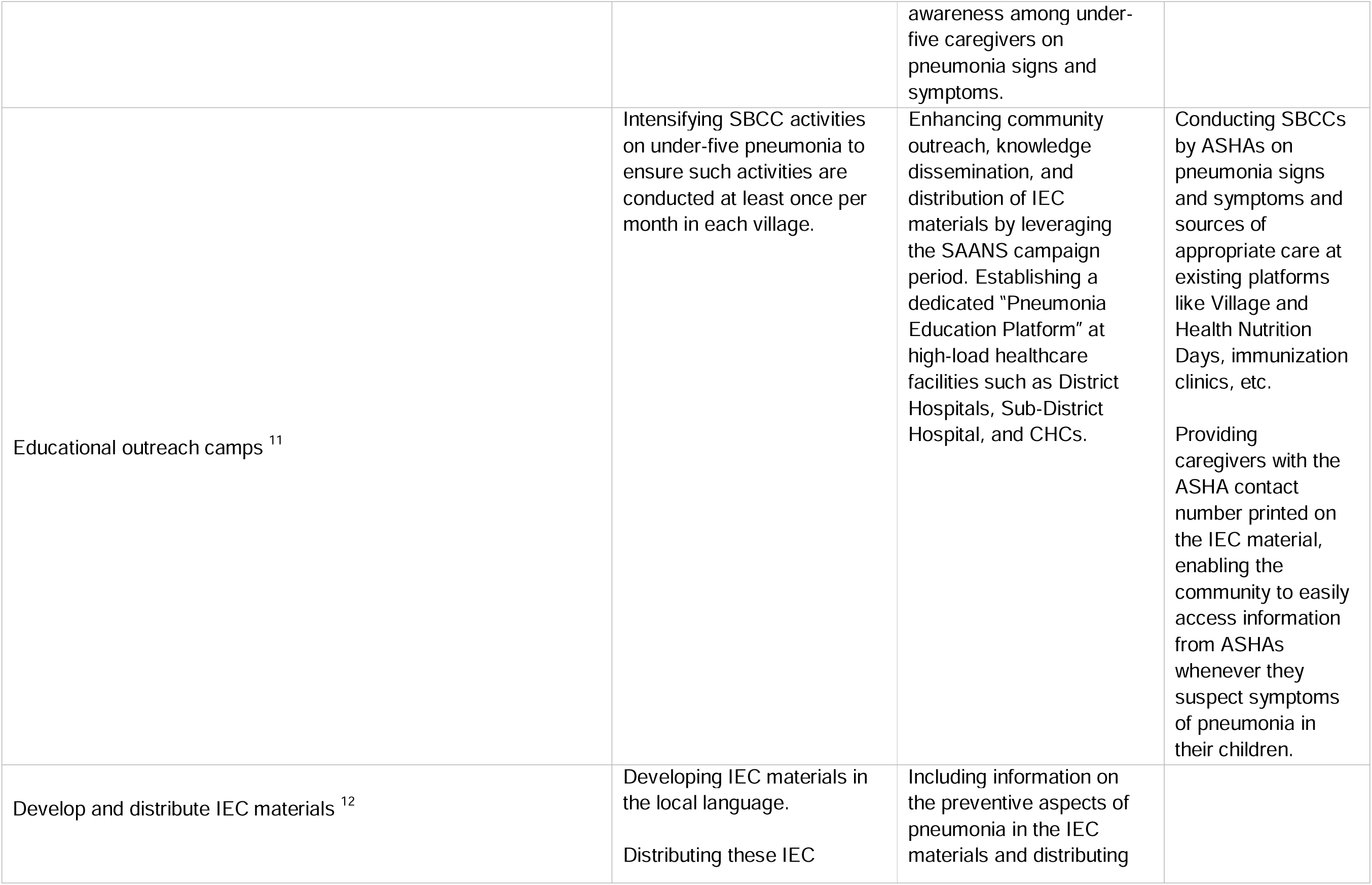
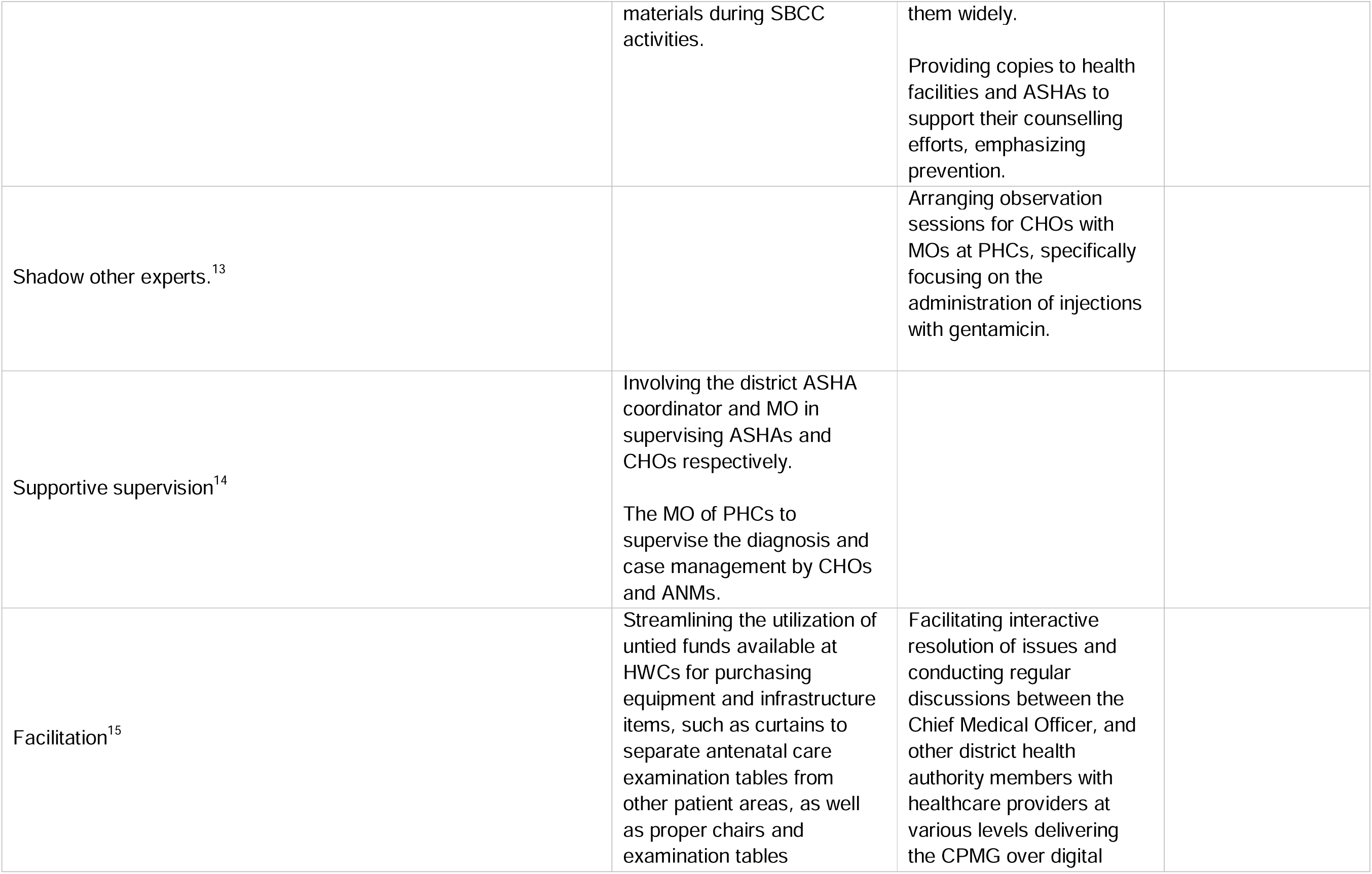
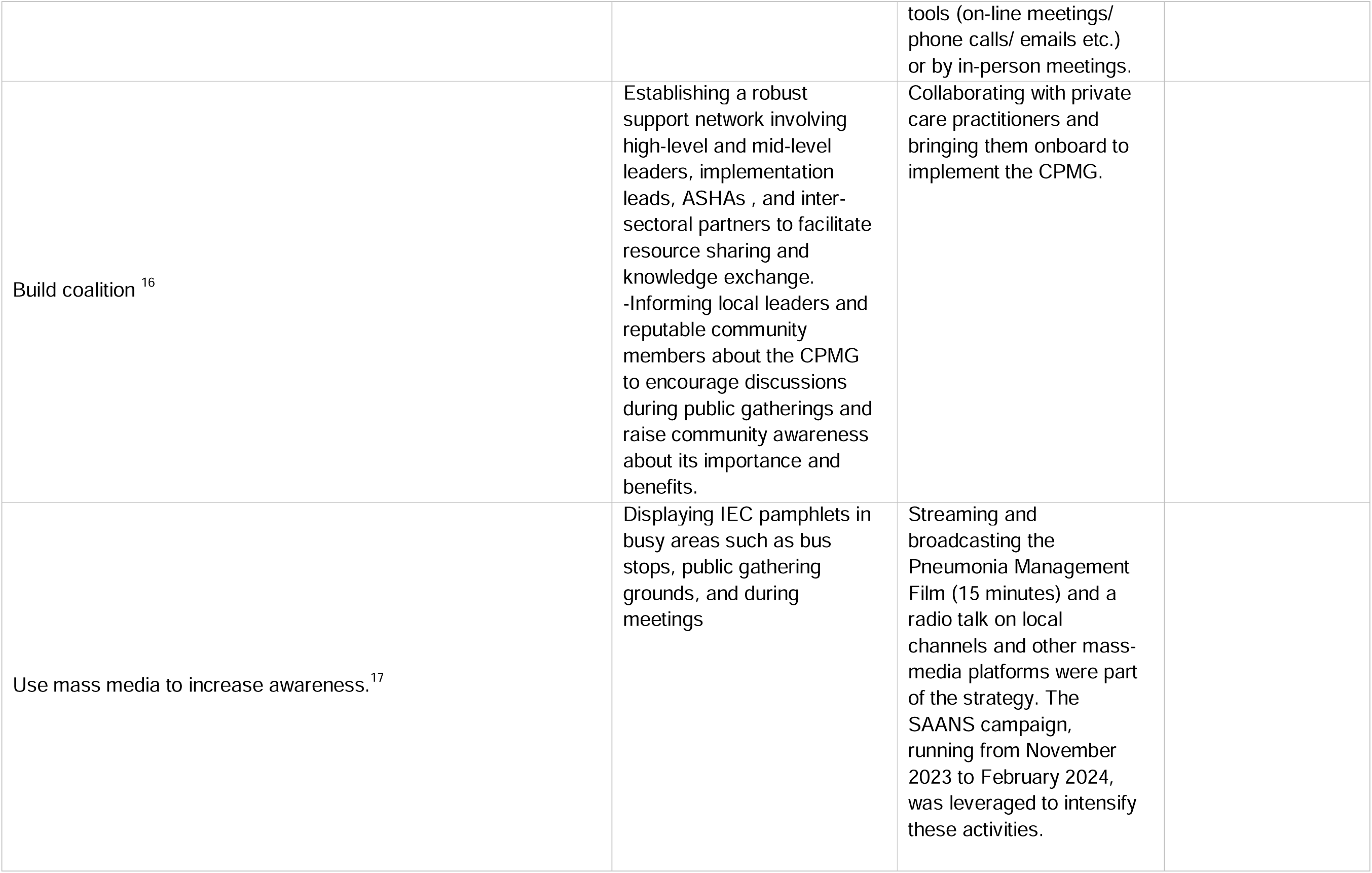
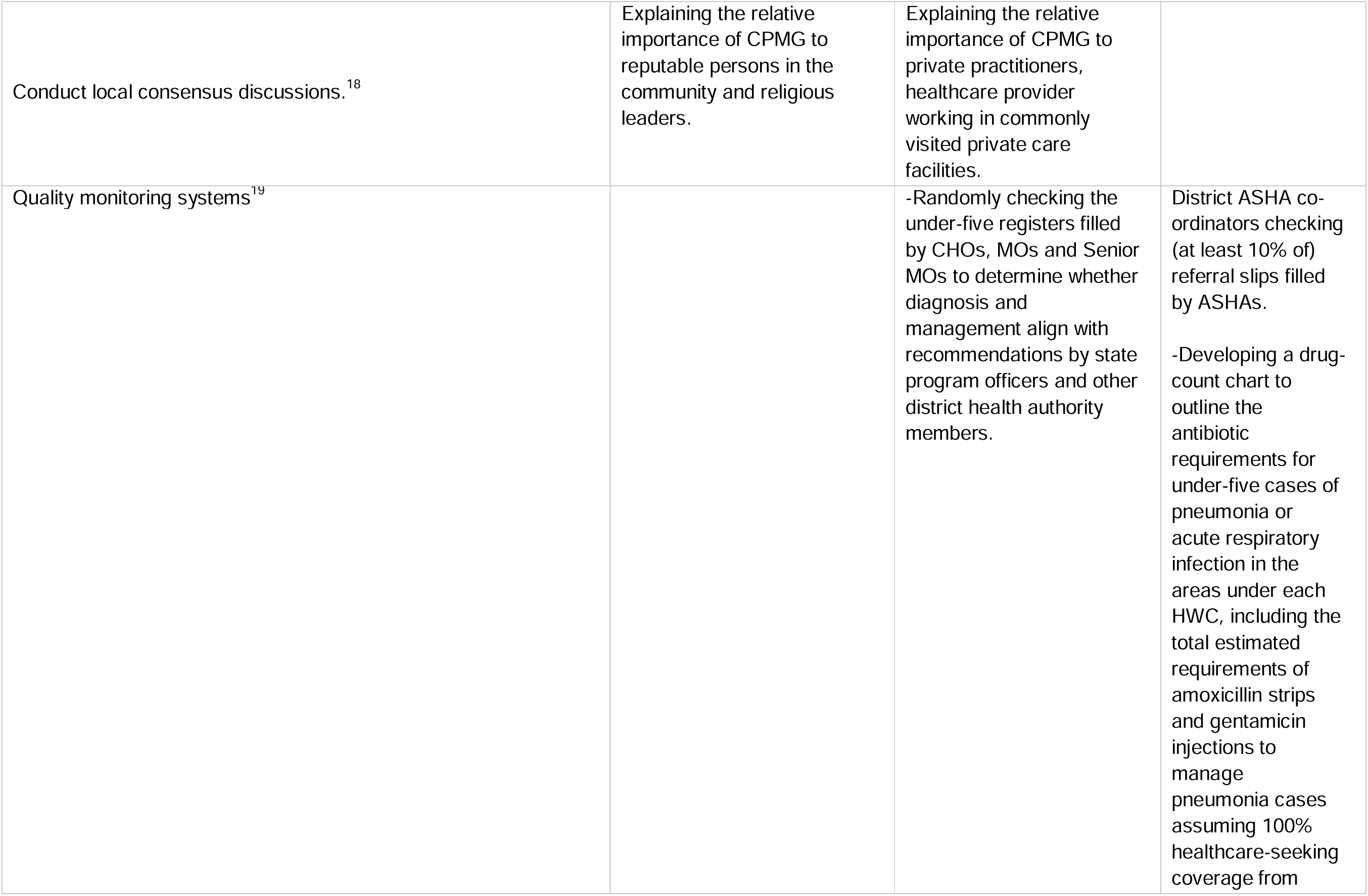
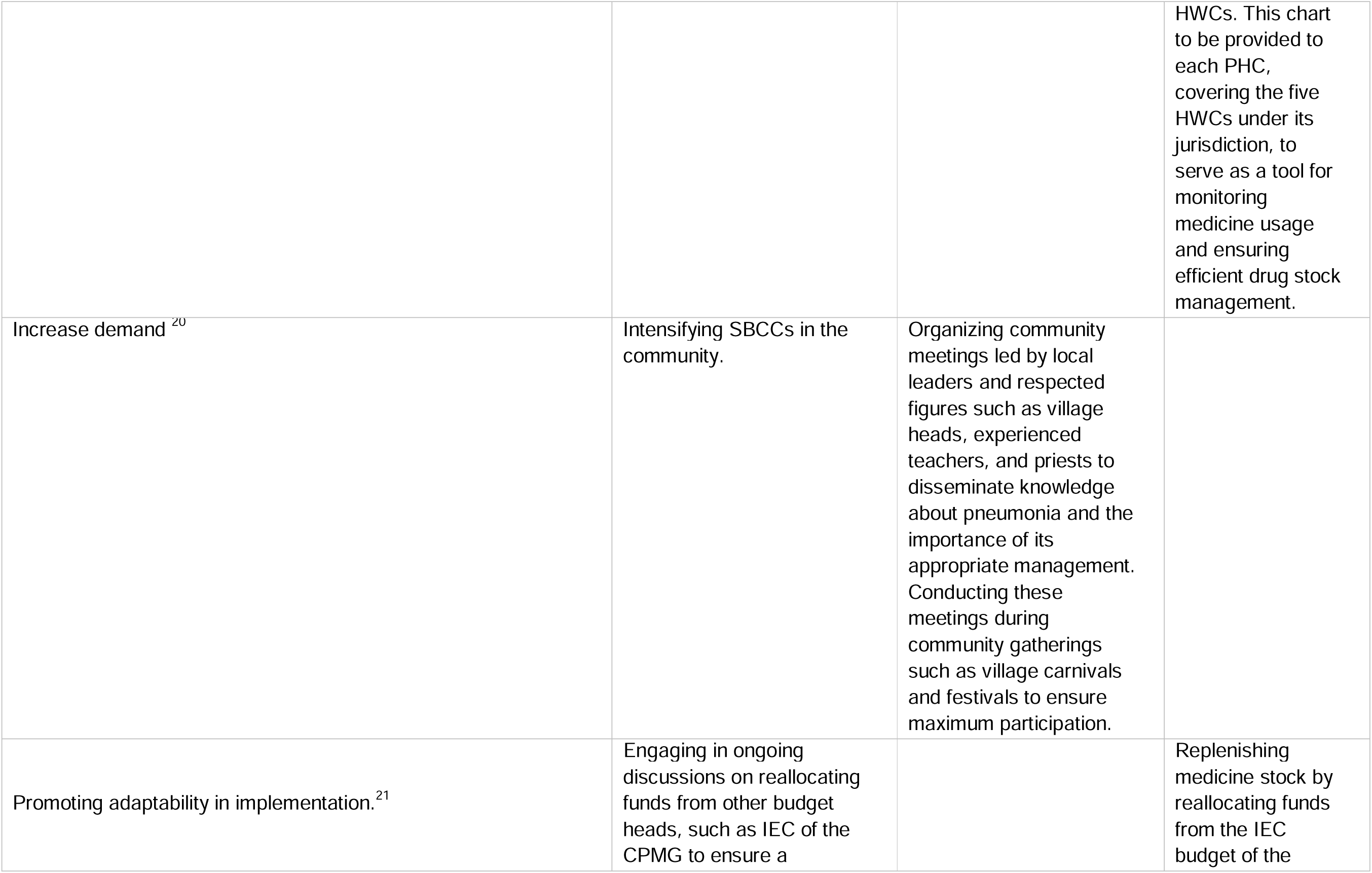
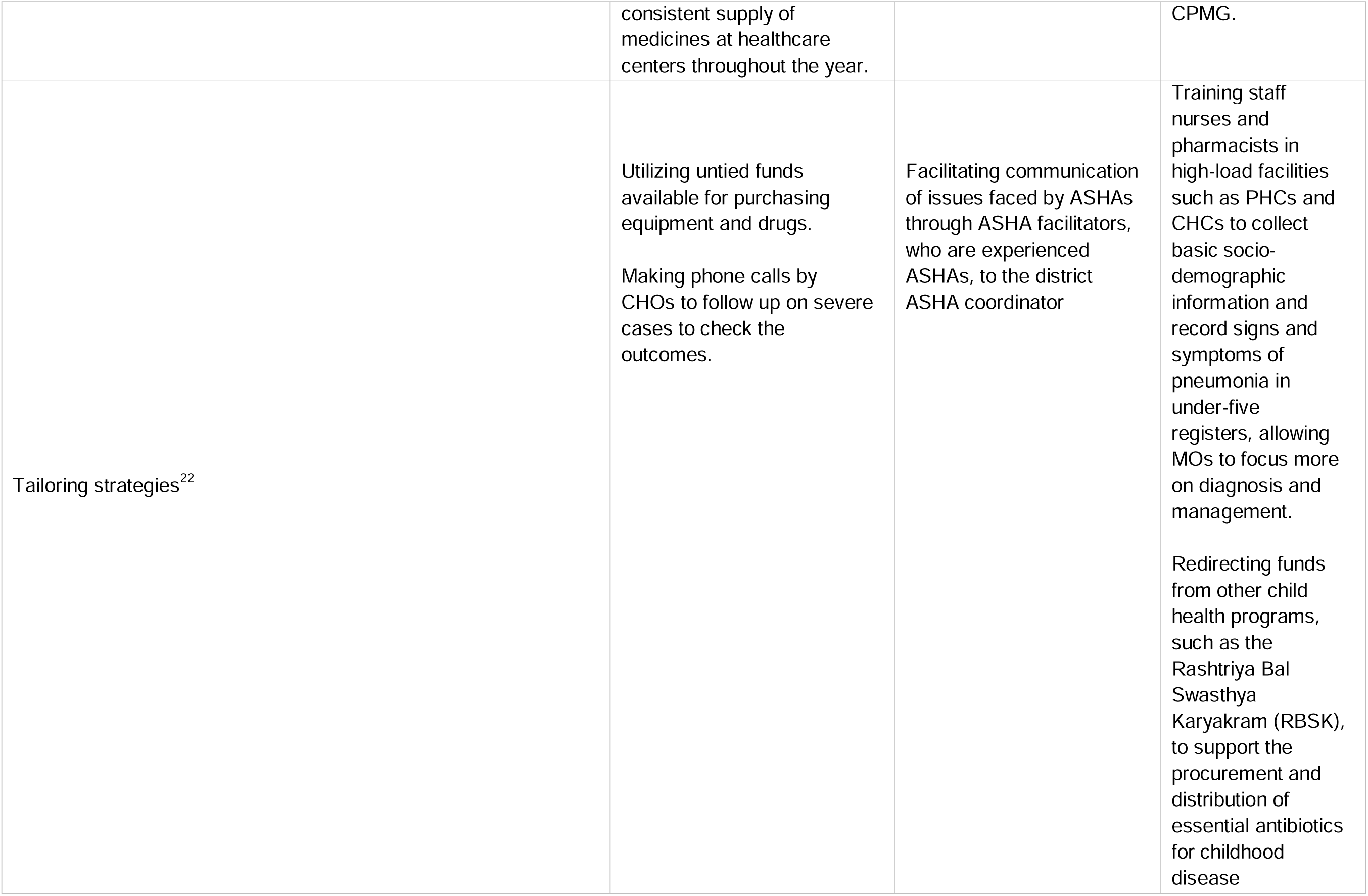
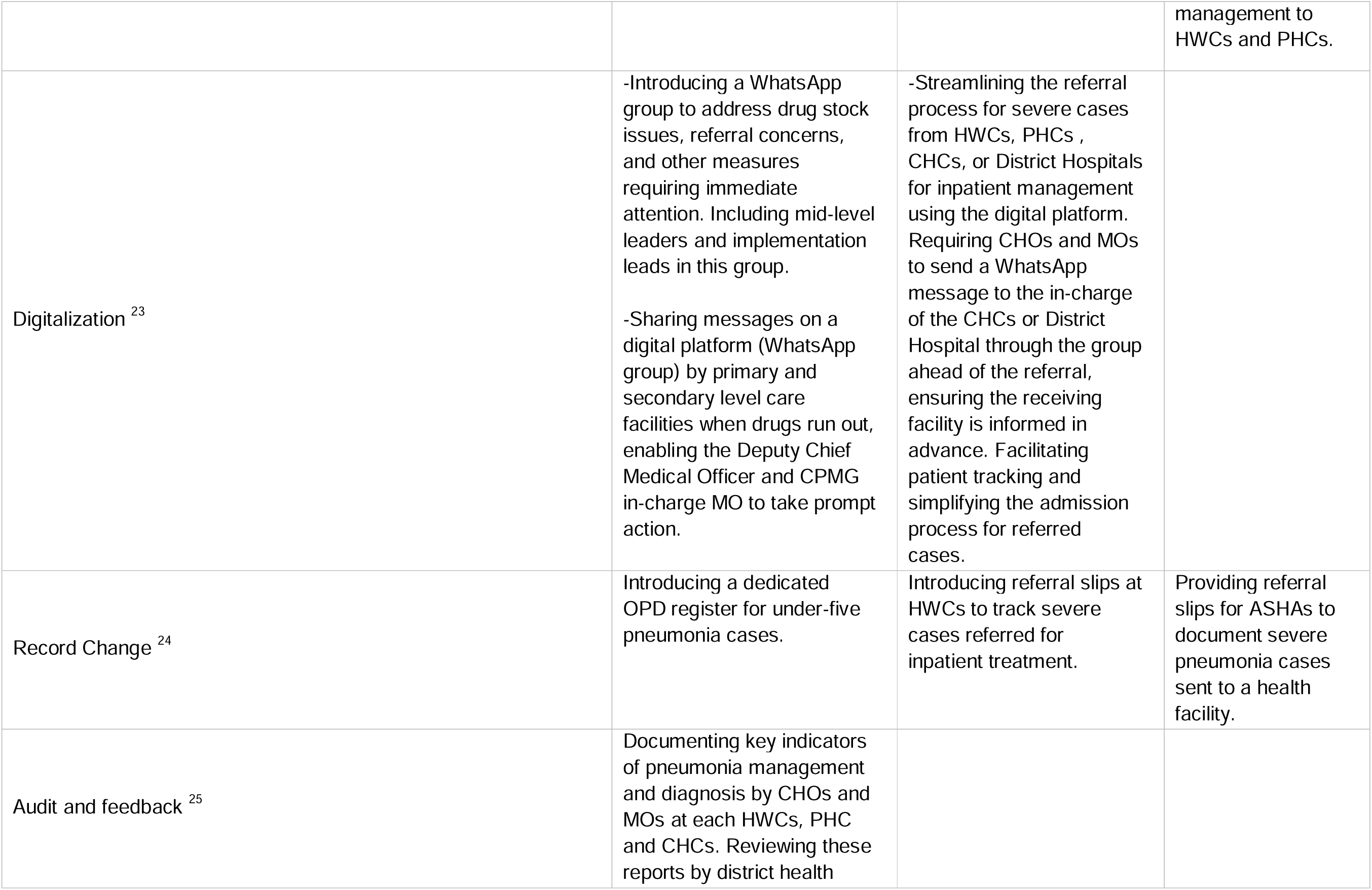
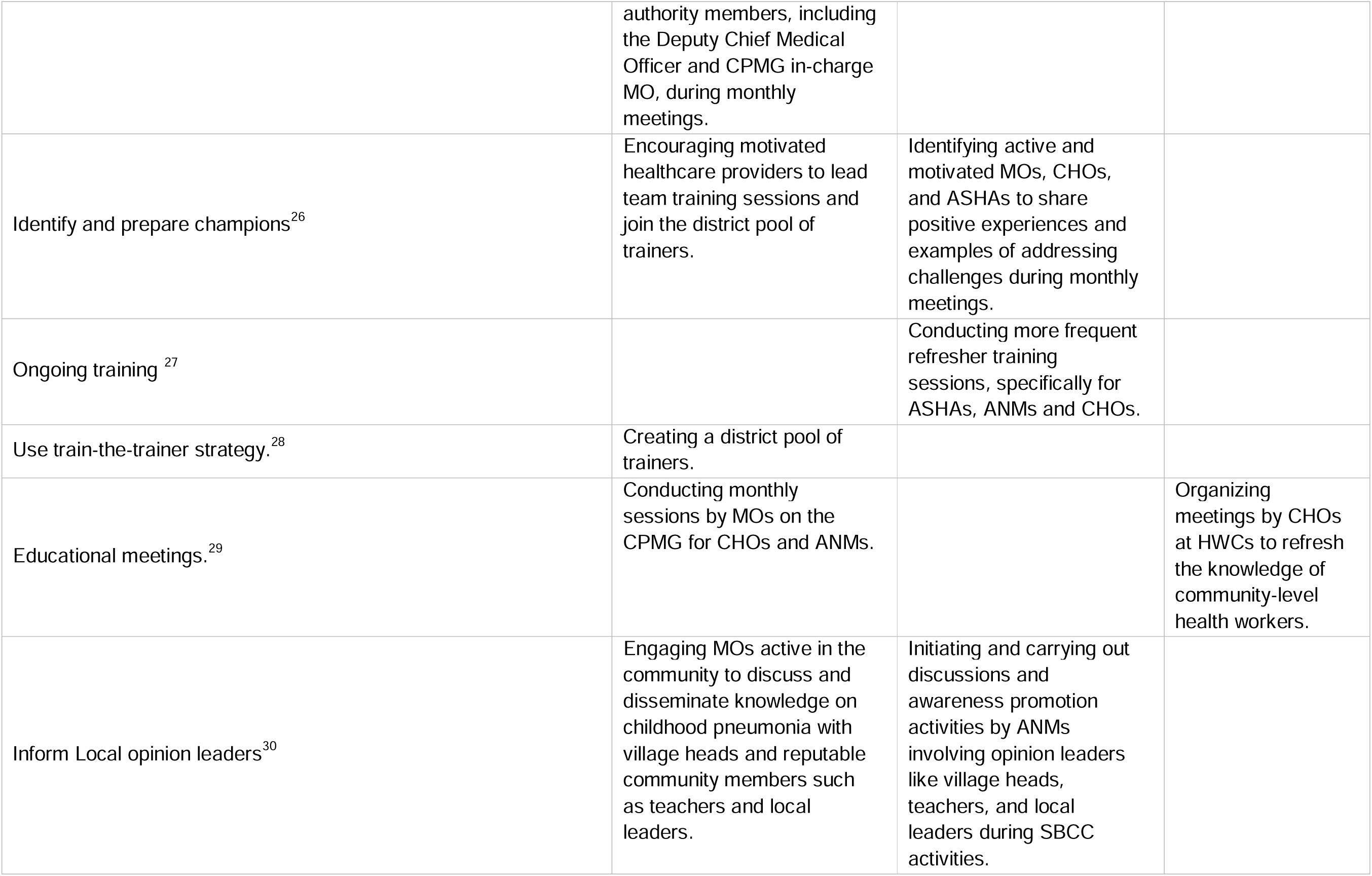
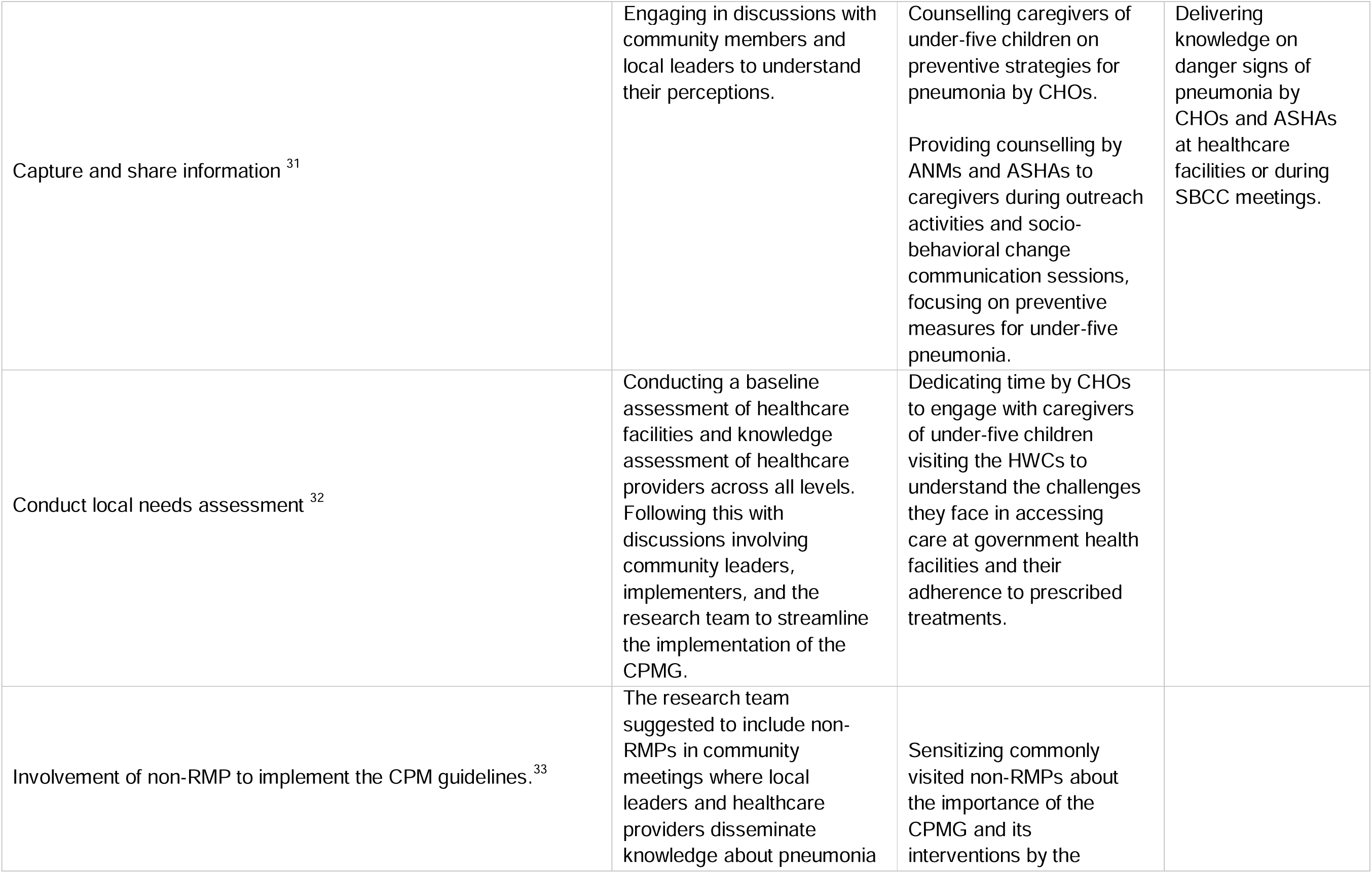
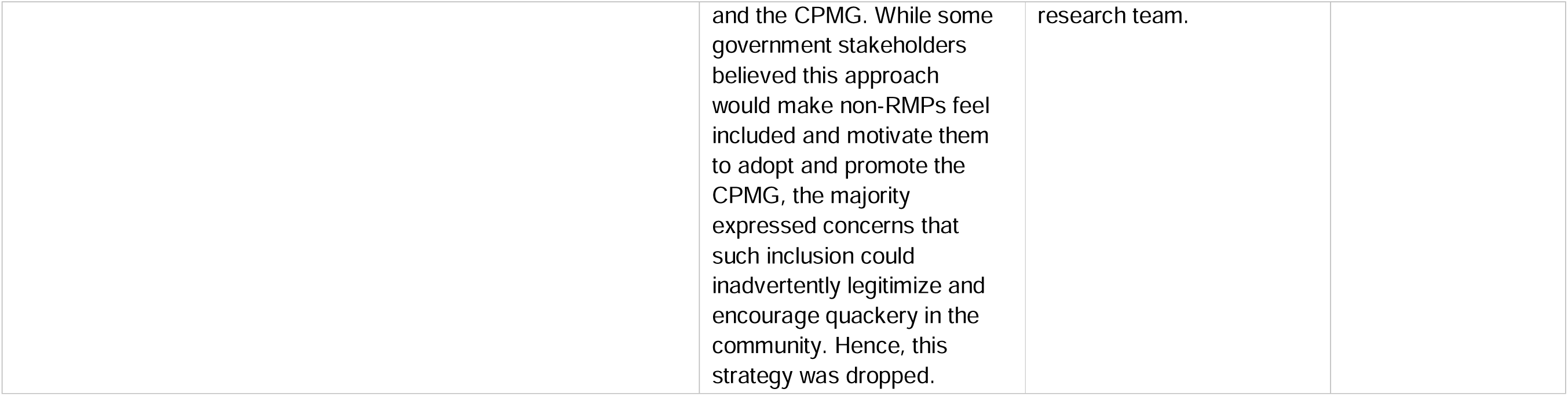
Co-designed implementation strategies with the stakeholders during the district and state review meetings and workshops.

**Table 4:**
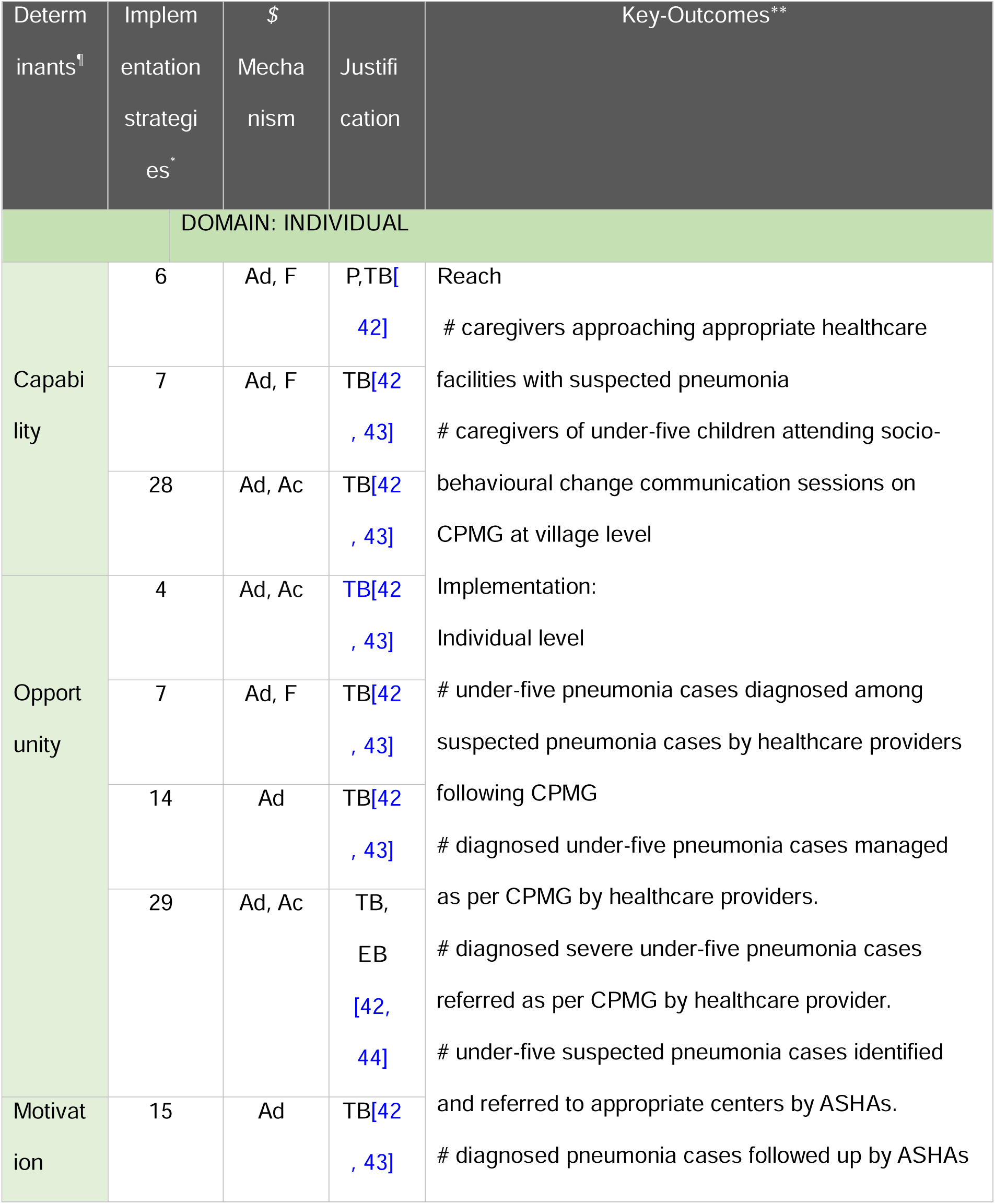
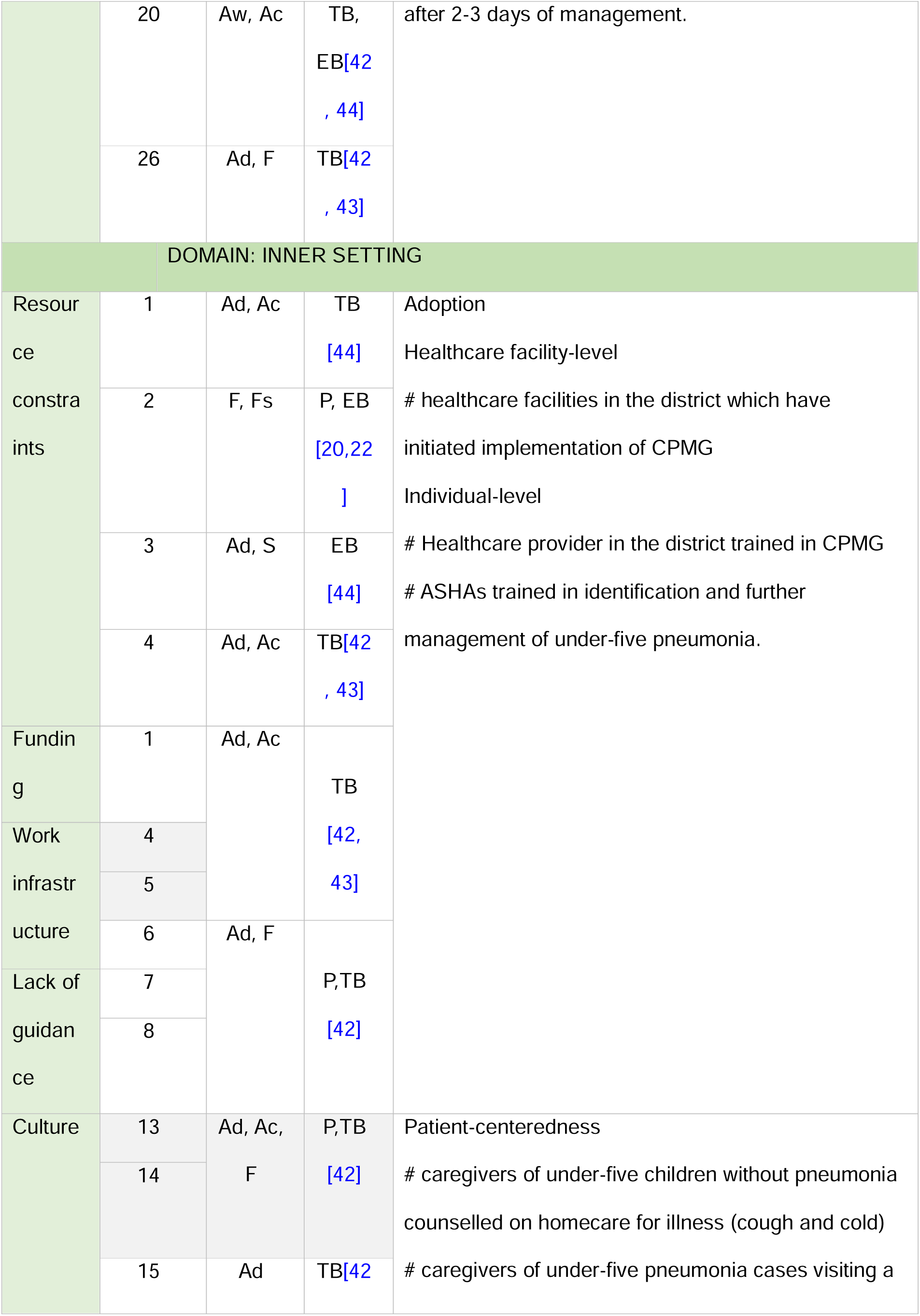
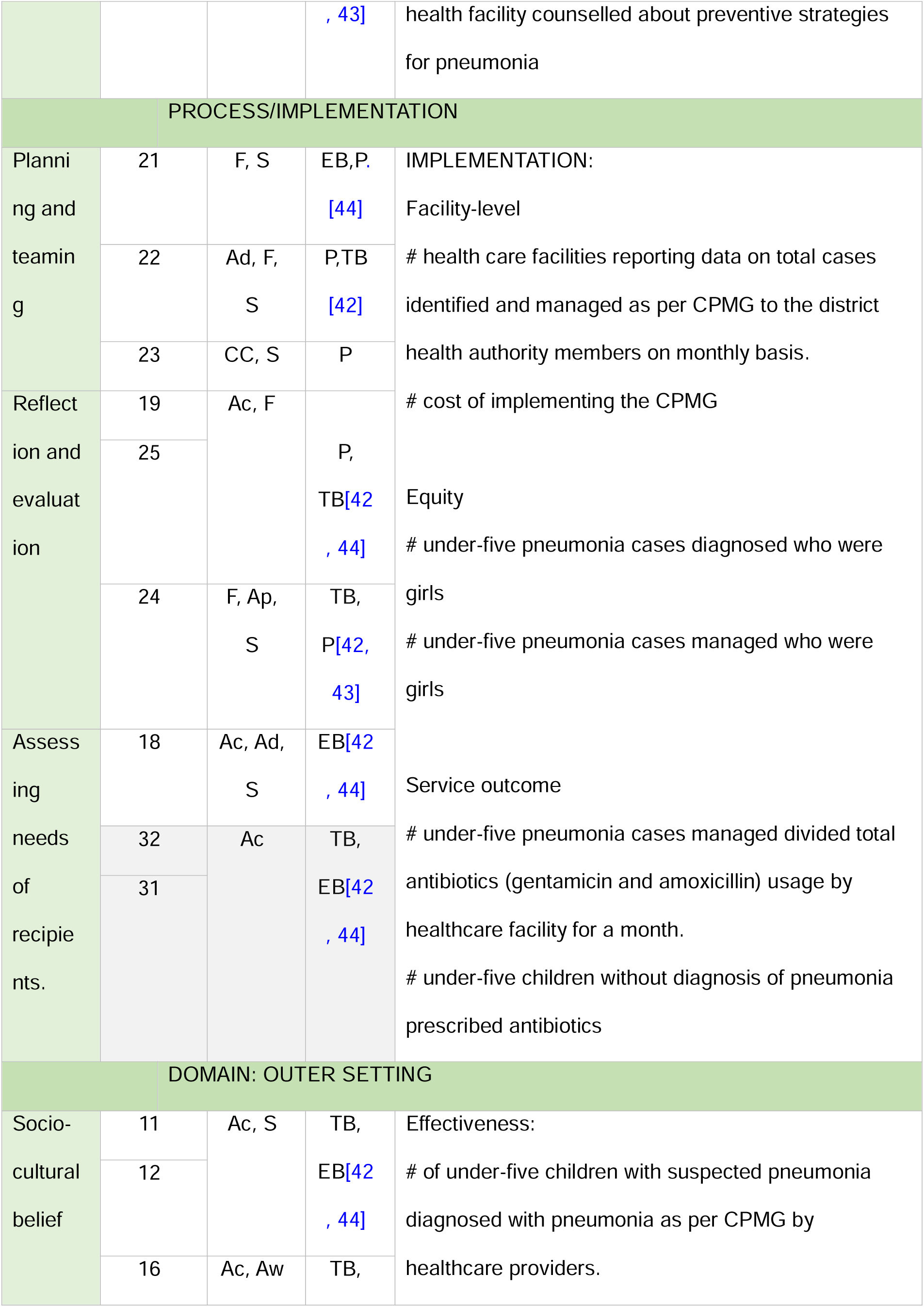
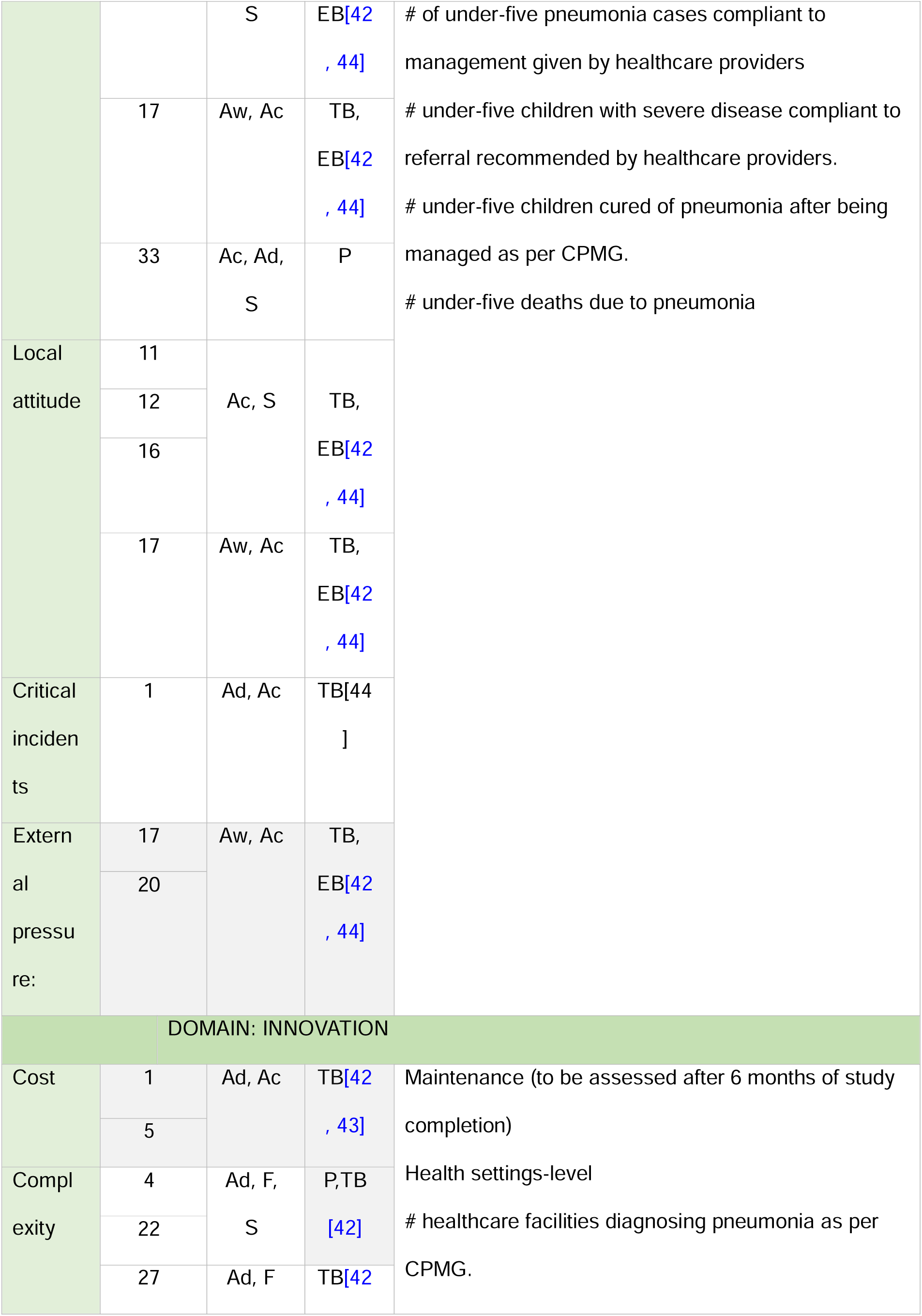
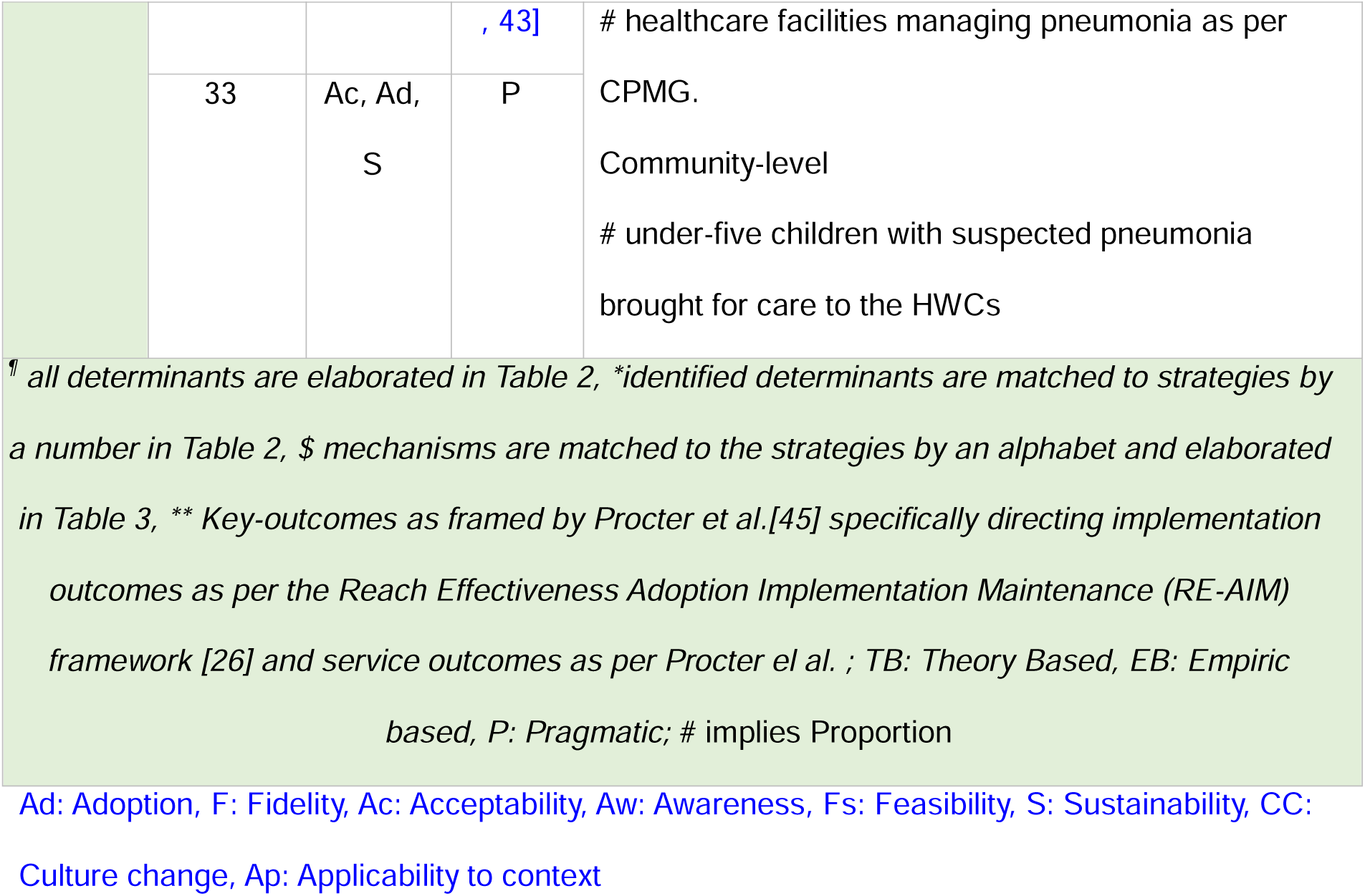
Implementation research Logic Model for achieving high coverage of appropriate management from primary-care settings among under-five children suffering from pneumonia.

For example, the ERIC strategy of obtaining formal commitment was operationalized through official directives from DHAm to implement the CPMG across all levels of care. These directives supported equipment procurement using the CPMG budget, regular review meetings, and improved accountability. Leadership engagement was reinforced through monthly indicator reviews and state-level advocacy for resource mobilization. Centralized procurement and distribution were refined to route drugs from the central warehouse to PHCs and HWCs, addressing earlier stockout issues. Network weaving involved regular meetings between DHAm, CHOs, and MOs to resolve operational gaps. Dynamic training strategies included creating a district trainer pool, hands-on CHO training, and private practitioner orientation. Early adopters were proposed to promote peer learning. Inclusion of private providers emerged as a critical strategy to address gaps in pneumonia care beyond the public system.

Other strategies focused on strengthening provider capacity and system-level implementation. Educational materials e.g. translated guidelines, facilitator guides, and visual IEC tools, were prioritized for accessibility across care levels. Simple reminders, like dosage charts in OPDs, supported clinical decision-making. Roles were adapted through coordination with the Integrated Child Development Scheme (ICDS), enabling Anganwadi workers to support ASHAs in pneumonia awareness, case identification, and referral tasks. Educational outreach was embedded into routine platforms like Village Health and Nutrition Days and intensified during the SAANS campaign. Clinical capacity was enhanced by enabling CHOs to shadow MOs from the PHCs during gentamicin administration and case history-taking. Supportive supervision mechanisms were institutionalized, with MO-in-Charges and ASHA coordinators overseeing case management practices. Community engagement improved through mass media campaigns and targeted IEC placement in high-traffic areas. Digital tools, such as WhatsApp groups, were introduced for real-time coordination of medicine stockouts and referrals. Finally, patient record systems were strengthened with dedicated under-five OPD registers and standardized referral slips, enabling better documentation and tracking of pneumonia cases across health system levels.

#### d. Mechanisms and justifications for implementation strategies

The assumed mechanisms of action and the justifications for the selections were recorded **(Table 4).** The final selected strategies aimed to improve the adoption, awareness of, acceptability, or the fidelity to the guidelines, or ultimately promoting the long-term sustainability of CPMG within routine primary care delivery. While the specific mechanisms were not formally documented, some indicative actions were observed. For example, obtaining formal commitment was assumed to increase acceptability and adoption of the CPMG, and we observed there was increased attention on pneumonia indicators in review meetings after DHAm issued formal directives related to CPMG implementation, and there were anecdotal instances where IEC funds were reallocated to support medicine procurement. Healthcare staff also reported more structured discussions during review meetings, which suggested a perception of increased performance oversight.

Three types of justifications were recorded: empirical evidence, theoretical constructs and practical experience. Empirical evidence indicated that strategies such as "Revise roles and responsibilities”, “Conduct ongoing training", and "Conduct educational meetings” could be helpful. Theoretical constructs and models, particularly those based on behavioural change principles, supported strategies such as "Identifying early adopters" and "Making training dynamic”. During the co-design sessions, healthcare professionals and implementation experts contributed practical insights from their own implementation experiences to justify why digitalization and involvement of non-registered medical practitioner (non-RMP) were likely to be helpful.

#### e. Development of an IRLM

By the end of Model 2, all three primary implementation outcome indicators exceeded the pre-specified 50% optimisation threshold: care-seeking reached 76.0% (193/254), appropriate diagnosis 92.7% (179/193), and fidelity to guideline-based management 86.0% (154/179), as presented in **Table 1** and **Figure 3**. Following the coding of notes, discussion points, and pragmatic suggestions from stakeholder convenings on the final optimized model, the IRLM was created, integrating additional complexities - mechanisms of change and justifications. The IRLM, presented in Table 3, provides a structured framework for strengthening primary care-based outpatient pneumonia management, while clarifying referral and support functions across higher-level facilities. Outcomes included measures such as the proportion of caregivers seeking care at appropriate facilities (reach), adherence to pneumonia management (effectiveness), the percentage of healthcare staff trained in CPMG, implementation initiation in healthcare facilities (implementation), and the percentage of facilities diagnosing pneumonia per CPMG six months post-IR (maintenance). Monthly tracking of these outcomes was expected to assess the effectiveness, adoption, and sustainability of the strategies, providing a replicable model for similar settings. The final stakeholder validation meeting was conducted in the first week of May 2024, with both in-person and virtual participation depending on the stakeholders’ preferences

## DISCUSSION

This study demonstrated how a structured, co-designed approach supported the adaptation and implementation of the CPMG within routine primary care delivery in a North Indian district. Utilizing the CFIR and ERIC frameworks, we systematically identified barriers, facilitators and relevant strategies and then refined the implementation strategies through iterative co-design workshops with government authorities and other stakeholders. A key contribution of this study is the development of the IRLM, which systematically links determinants, strategies, and expected outcomes, facilitating transparent strategy selection for optimized primary care-based pneumonia management.

Most of the ERIC strategies required only minimal modifications, and only a few needed substantial contextual tailoring. For example, train-the-trainer models, expert shadowing, and structured educational outreach were quickly integrated into existing healthcare training structures, making them highly feasible. The district-level trainer pool and peer-group discussions ensured sustained capacity-building and knowledge retention and made structured refresher training feasible within the government health systems. Similarly, digital tools for real-time coordination and referral tracking were well-received by stakeholders. These tools supported decision-making, documentation, and continuity of care within primary care OPD workflows, and were strengthened with a dedicated platform to coordinate stockouts and referral processes, an approach shown to improve healthcare delivery in LMICs [34] Dynamic training models, and utilization of digital health technology have also been found to be effective in managing childhood diseases in similar contexts.[35-37]

Feasibility and acceptability of strategies were further improved through stakeholder-driven modifications during implementation. For instance, demand-generation strategies initially proposed as broad community engagement interventions were refined into mass-media campaigns, targeted IEC placement, and leveraging the SAANS campaign period for pneumonia awareness. These stakeholder-driven adaptations helped integrate pneumonia education within routine healthcare activities e.g., Village Health and Nutrition Days, to avoid overburdening frontline health workers, suggesting potential for sustainability within existing service delivery structures. The study also indicates that inter-sectoral collaboration and task-shifting, particularly the inclusion of Anganwadi workers in pneumonia awareness and case identification, was feasible and scalable.[38],[39]

The integration of private healthcare providers into the CPMG training was another key adaptation that enhanced the feasibility and broadened the scope of district efforts to improve pneumonia management. Engaging private practitioners complemented primary care services by improving referral coordination and data completeness, demonstrating potential for sustainable public-private partnerships. This aligns with findings from Nepal, where similar cross-sectoral partnerships improved adherence to national treatment guidelines for tuberculosis.[40]

Strong leadership engagement and structured review mechanisms were essential for ensuring effective strategy adaptation and sustainability within primary care-oriented health systems. Regular performance reviews and supervisory feedback loops helped maintain implementation momentum, reinforcing accountability mechanisms that are often weak in large-scale health programs. Stakeholder-driven reallocation of resources, such as redirecting IEC funds to medicine procurement, illustrated how flexible financial planning enhances program implementation. Previous studies have also highlighted leadership as a key determinant of successful implementation of interventions in a low-resource setting.[41]

This project addressed both demand- and supply-side factors. Key supply-side improvements included structured training programs, enhanced record-keeping, streamlined medicine procurement, and supportive supervision. On the demand side, mass media campaigns, targeted caregiver counselling, and community outreach increased awareness and timely care-seeking. This comprehensive approach strengthened both healthcare system capacity and community engagement, were associated with improvements in pneumonia management indicators. Overall, the participatory co-design approach contributed to developing a contextually feasible implementation model with potential for scale, suggesting potential suitability for wider adoption within similar settings across India.

The IRLM functions as an adaptive tool rather than a static framework because it allows for ongoing modifications based on real-time feedback, emerging data, and stakeholder input. This adaptability is essential in dynamic healthcare environments, where local contexts and barriers can change. By supporting continuous learning and iterative refinement, the IRLM ensures that implementation strategies remain relevant, feasible, and responsive to evolving challenges. One important limitation of the IRLM as developed in this study should be acknowledged. Although the IRLM is conceptually designed to document the mechanisms by which implementation strategies are expected to produce change, that is, the explanatory pathway linking strategy to outcome, the Results section notes that specific mechanisms were not formally documented during Phase II model iterations. This occurred because the primary focus during Phase II was on iterative optimisation and strategy refinement in real time, rather than on mechanism elicitation, and formal mechanism documentation was deferred to the final IRLM developed at the end of Phase II. As a result, the mechanisms presented in Table 4 reflect the research teams and stakeholders’ assumed pathways, grounded in theory and practical experience, rather than empirically verified mechanisms of change. We acknowledge this as a limitation and note that formal mechanism evaluation, including testing whether strategies worked through their hypothesised pathways, is planned as part of the Phase III evaluation. Future studies using the IRLM should consider incorporating structured mechanism elicitation, for example through theory of change workshops or contribution analysis, as an explicit step in the co-design process rather than a post-hoc addition.

### Strengths and Limitations

To the best of the authors’ knowledge, this is among the few studies from low-resource Asian settings to report findings using a structured application of implementation research frameworks, specifically CFIR, ERIC, and the IRLM, to develop and refine context-specific strategies for childhood pneumonia management in primary care. The CFIR-ERIC Barrier Busting Tool strengthened strategy selection, ensuring strategies were grounded in expert recommendations yet adaptable to local constraints. The creation of the IRLM was based on a systematic approach to aligning implementation strategies with identified barriers, mechanisms of change, and measurable outcomes, ensuring transparency, rigour, and reproducibility. The model incorporated key indicators using the RE-AIM framework, which will support monitoring during scale-up. The participatory co-design approach, engaging government stakeholders, frontline providers, ASHAs, caregivers, and PRI representatives, strengthened contextual relevance and ownership, which are critical determinants of sustainability. Although the model was tailored to Palwal’s local context, the structured, theory-driven, and stepwise procedure offers a replicable process for other LMIC settings rather than promoting fixed strategies.

Several important limitations should be acknowledged. First, the study did not include a control or comparison group. The pre-post optimisation design within the learning cluster means that observed improvements in care-seeking, diagnosis, and management cannot be attributed solely to the implementation strategies; secular trends, the Hawthorne effect arising from the research team’s continuous presence, and concurrent contextual changes, such as the post-COVID recovery in care-seeking and the SAANS campaign period, may have contributed to observed improvements. Causal inference is therefore not possible from this phase of the study, and outcome figures should be interpreted as optimisation-phase indicators rather than as evidence of effectiveness. No formal statistical testing was conducted to assess whether observed improvements exceed expected random variation, and secular trends, the Hawthorne effect, and concurrent contextual changes cannot be statistically excluded as explanations for the observed improvements. The quasi-experimental evaluation of the optimised model across the full study area, which will permit stronger causal inference through comparison with a broader baseline and concurrent monitoring, will be reported in the Phase III paper.

Second, the generalisability of findings from the learning cluster to the broader district and to other similar districts requires careful consideration. The learning cluster was selected purposively to represent geographic and facility-level diversity within Palwal, but it covered approximately 50,000 of the district’s 1,285,000 inhabitants and four of its 72 HWCs. The cluster’s facilities and populations may not fully represent the heterogeneity of the district, particularly in more remote or underserved areas. Furthermore, Palwal’s specific combination of infrastructure constraints, workforce composition, and community attitudes may not be directly transferable to other districts or states in India or to other LMIC contexts. The structured CFIR-ERIC-IRLM approach is intended to be replicable, but the specific strategies identified will require local adaptation and re-contextualisation in other settings.

Third, the study relied heavily on routine programme records, particularly under-five OPD registers, as the primary source of quantitative outcome data, which introduces a significant risk of measurement bias. The paper itself documents substantial data quality problems at baseline, including absent or poorly maintained OPD registers, incomplete entries, and facilities prescribing antibiotics without recording a confirmed diagnosis. While the study introduced standardised data extraction procedures and fortnightly quality checks to mitigate this risk, the validity of indicator trends, particularly in the early model periods, remains dependent on the quality of records that were, at baseline, acknowledged to be suboptimal. This should be borne in mind when interpreting the optimisation indicator figures. The OPD register validation procedures introduced in Phase II are described in the Methods and represent a partial mitigation; however, residual measurement bias cannot be excluded.

Fourth, the qualitative component relied on purposive sampling, which may not have fully captured the perspectives of less vocal or more marginalised caregivers, particularly those from very low socioeconomic backgrounds or remote villages, who may face distinct barriers to care-seeking. Social desirability bias may have influenced healthcare provider responses, particularly given the research team’s supervisory role in the learning cluster. Fifth, the research team’s active involvement in model implementation creates the potential for performance bias, as the presence of trained observers and the intensity of research team engagement during Phase II are unlikely to be replicable at scale without dedicated implementation support. This limitation is acknowledged as a key rationale for Phase III, which evaluates the model under conditions closer to routine government-led implementation.

## Conclusion

This study presents a systematic, participatory, and theory-driven approach to developing and optimising a primary care-based implementation model for childhood pneumonia management in a resource-constrained district of North India. Through the structured application of CFIR, ERIC, and the IRLM, and iterative co-design with government and community stakeholders, a contextually feasible and locally owned set of implementation strategies was developed and refined across three successive model iterations. The optimisation process demonstrated progressive improvements in care-seeking, diagnosis, and guideline-concordant management within the learning cluster, providing a foundation for the district-wide scale-up reported separately. The co-designed IRLM offers a replicable, adaptive framework for translating evidence-based guidelines into primary care practice in similar LMIC settings, not as a fixed prescription, but as a flexible, evidence-informed process that can be contextualised to local barriers and health system structures. Findings from the Phase III district-wide implementation and evaluation will be reported in a forthcoming publication.

## Declarations

### Ethics approval and consent to participate

Ethical approval was granted by the ethical committees of the Society for Applied Studies (SAS/ERC/IR Pneumonia/2021), the Regional Ethics Committee of Western Norway (2022/531608) and the World Health Organisation (WHO/ ERC.0003652). Additionally, this study has obtained the government of Haryana state (Memo no. HSHRC/2022/505) and Health ministry steering committee (approval date: 19 Dec 2022, proposal id 2022-17596) approvals. Written informed consent was obtained from all healthcare providers, community health workers, and caregivers prior to participation, after the study information sheet was read out and explained to them in their preferred language. Participants were informed of their right to withdraw or request to stop recording the interviews at any time without consequence. This study was conducted in accordance with the principles of the Declaration of Helsinki and relevant national ethical guidelines.

### Consent for publication

Not applicable.

### Availability of data and materials

The data used and/or analysed during the current study are available from the primary author/s and corresponding author Barsha Gadapani Pathak and/or and Sarmila Mazumder upon reasonable request.

### Authors’ contributions

SM and BGP conceived the study. SM, BGP, and TM coordinated the study at the study site. BGP was involved in data collection, and analysis and wrote initial and subsequent drafts of manuscripts. AB, MS and MH assisted in data collection and analysis of data. SM and IFS provided input into the analysis, reviewed the preliminary results, and assisted in manuscript drafting. RS, SB, and VY provided support in data collection and oversaw the project. YN provided necessary resources for the study and study oversight (annual reporting). All authors read, critically reviewed, and approved the final manuscript.

## Acknowledgements

The authors are also deeply grateful to Dr. Vinod Kumar Anand for serving as the Master Trainer and leading the training of the study team and government officials in childhood pneumonia management guidelines. Authors extend their appreciation to the Chief Medical officer, Palwal and other healthcare providers and community health workers for their active involvement. Special thanks are due to Dr. Naveen Garg (Deputy CMO), Palwal District Hospital, for his invaluable support and facilitation throughout the implementation of the project. The authors express their sincere gratitude to all caregivers of under-five children who participated in this study and generously shared their experiences. Finally, the authors acknowledge the unwavering efforts and commitment of the study staff and data management team, whose dedication was instrumental to the successful execution of this research.

## Abbreviation

ANM: Auxiliary Nurse Midwife
ASHA: Accredited Social Health Activist
CFIR: Consolidated Framework for Implementation Research
CHC: Community Health Centre
CHO: Community Health Officer
CPMG: Childhood Pneumonia Management Guidelines
COREQ: Consolidated Criteria for Reporting Qualitative Research
DH: District Hospital
DHAm: District Health Authority members
ERIC: Expert Recommendations for Implementing Change
HWC: Health and Wellness Centre
ICDS: Integrated Child Development Scheme
IDI: In-depth interview
IEC: Information, Education and Communication
IR: Implementation Research
IRLM: Implementation Research Logic Model
LMIC: Low- and middle-income country
MO: Medical Officer
Non-RMP: Non-Registered Medical Practitioner
OPD: Outpatient Department
PHC: Primary Health Centre
PRI: Panchayati Raj Institution
RE-AIM: Reach, Effectiveness, Adoption, Implementation, Maintenance
SAANS: Social Awareness and Action to Neutralise Pneumonia Successfully
SBCC: Social and Behavioural Change Communication
SHAm: State Health Authority members
SMO: Senior Medical Officer
StaRI: Standards for Reporting Implementation Studies

